## Supplemental Figures and Tables for "Genetic regulation of serum IgA levels and susceptibility to common immune, infectious, kidney, and cardio-metabolic traits"

*Lili Liu et al.*

### SUPPLEMENTAL FIGURES

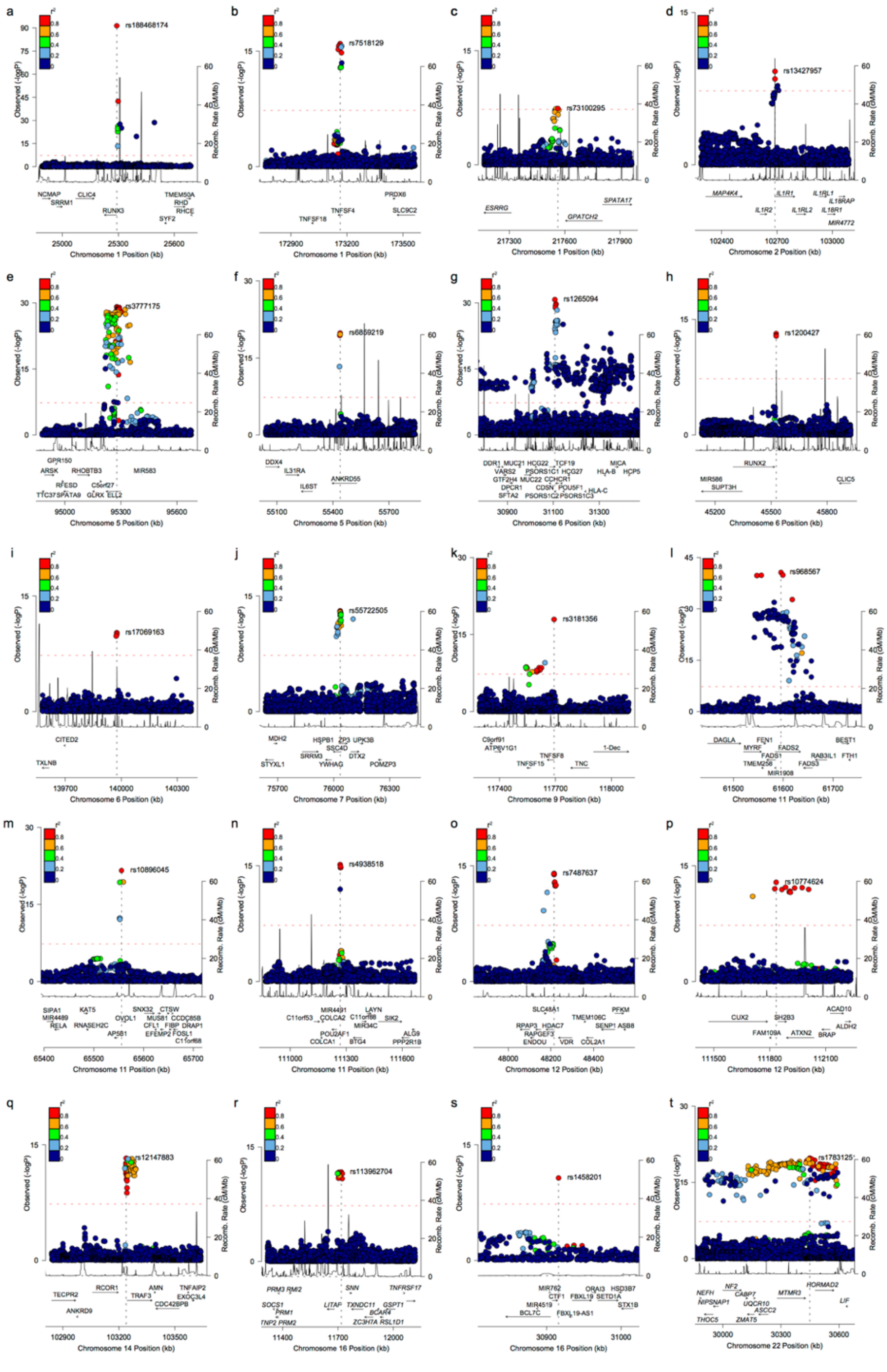

**Supplementary Figure 1. Regional plots for 20 genome-wide significant loci: (a)** *RUNX3*, **(b)** *TNFSF4*, **(c)** *GPATCH2*, **(d)** *IL1R1*, **(e)** *ELL2*, **(f)** *ANKRD55*, **(g)** *HLA*, **(h)** *RUNX2*, **(i)** *CITED2*, **(j)** *ZP3/SSC4D*, **(k)** *TNFSF8/TNFSF15*, **(l)** *FADS1/TMEM258*, **(m)** *OVOL1/RELA*, **(n)** *POU2AF1*, **(o)** *HDAC7/VDR*, **(p)** *SH2B3*, **(q)** *TRAF3*, **(r)** *LITAF*, **(s)** *CTF1*, and **(t)** *HORMAD2/LIF* loci. The x-axis shows the physical position in Mb (hg19 coordinates) along with known genes in the region; the left y-axis presents -log10 p-values for association statistics and the right y-axis shows the recombination rate across the region; the dotted horizontal line indicates a genome-wide significant threshold of 5.0E-08.

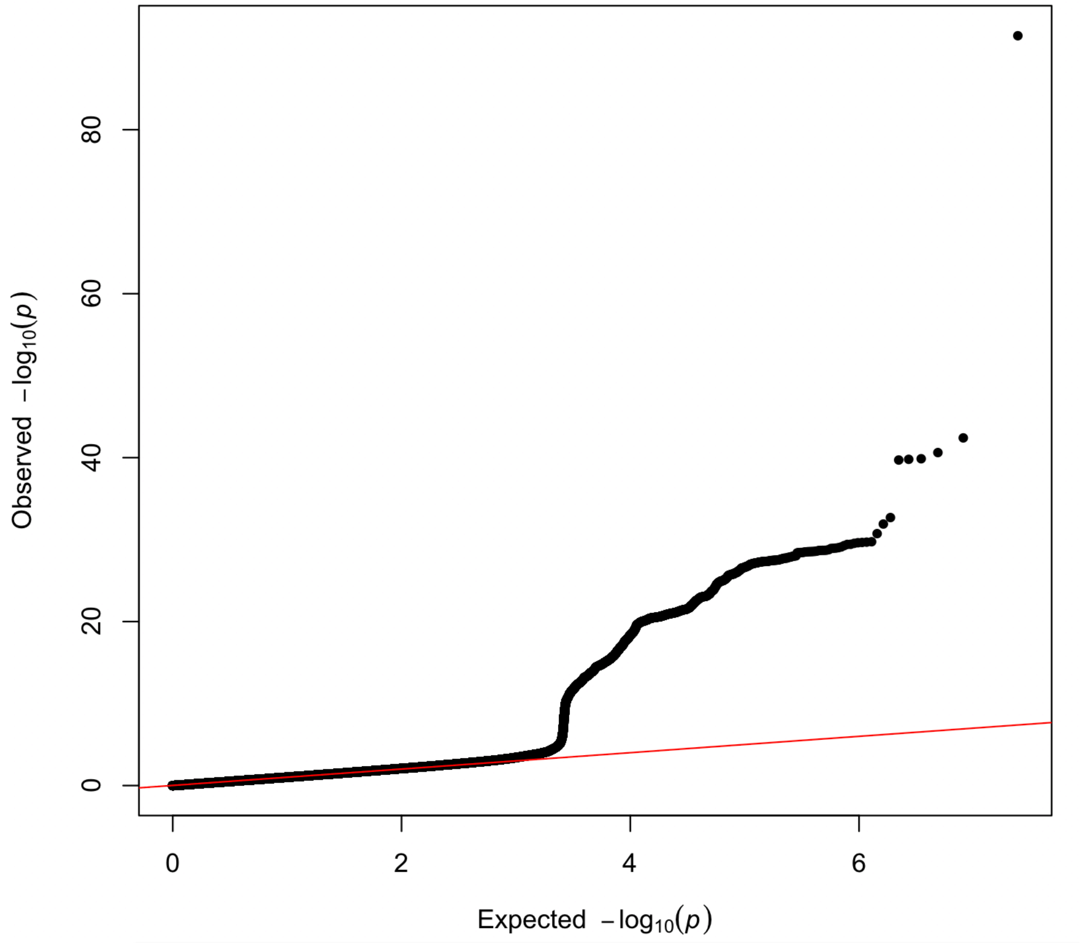

**Supplementary Figure 2. Quantile-quantile (QQ) plot of the trans-ethnic meta-analysis (total N = 41,263 individuals).** The overall meta-analysis genomic inflation factor ($\lambda$) was estimated at 1.016.

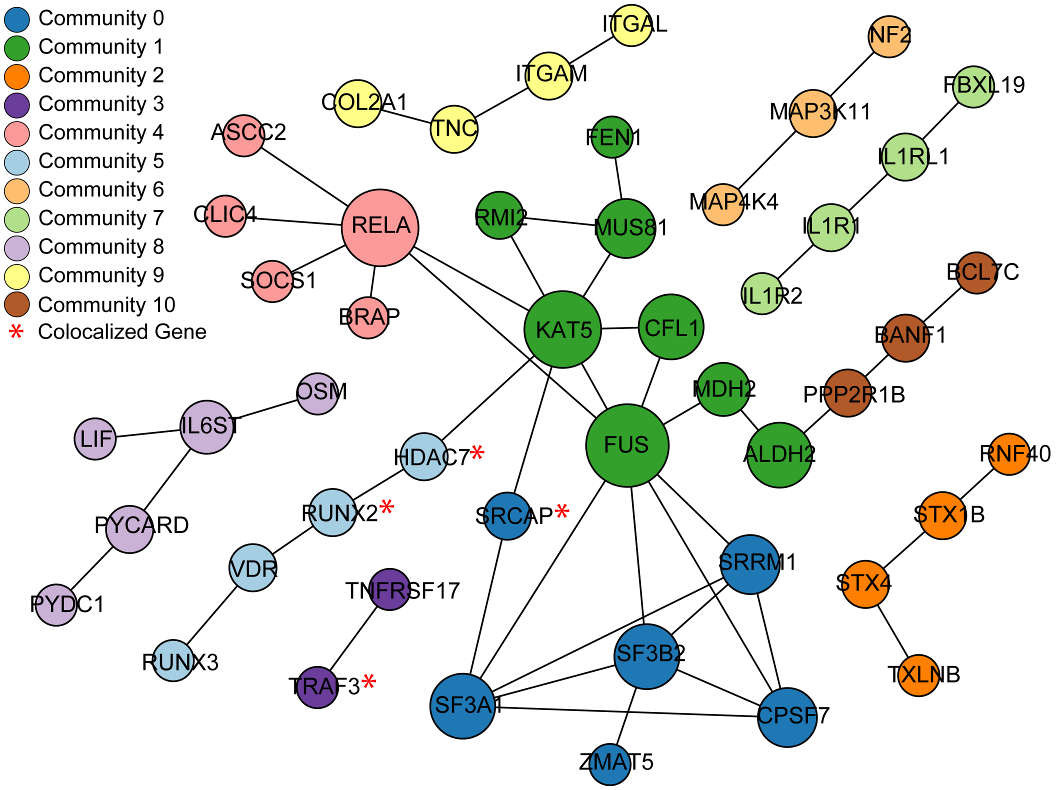

**Supplementary Figure 3. Protein-protein interaction (PPI) network for positional candidate genes at GWAS loci:** each color represents a distinct network module (a group of inter-connected genes); red asterisk indicates e-genes co-localized with GWAS loci. Overall this PPI network has more connectivity than expected by chance (permutation P<2e-03). The PPI information was obtained from InWeb_IM.

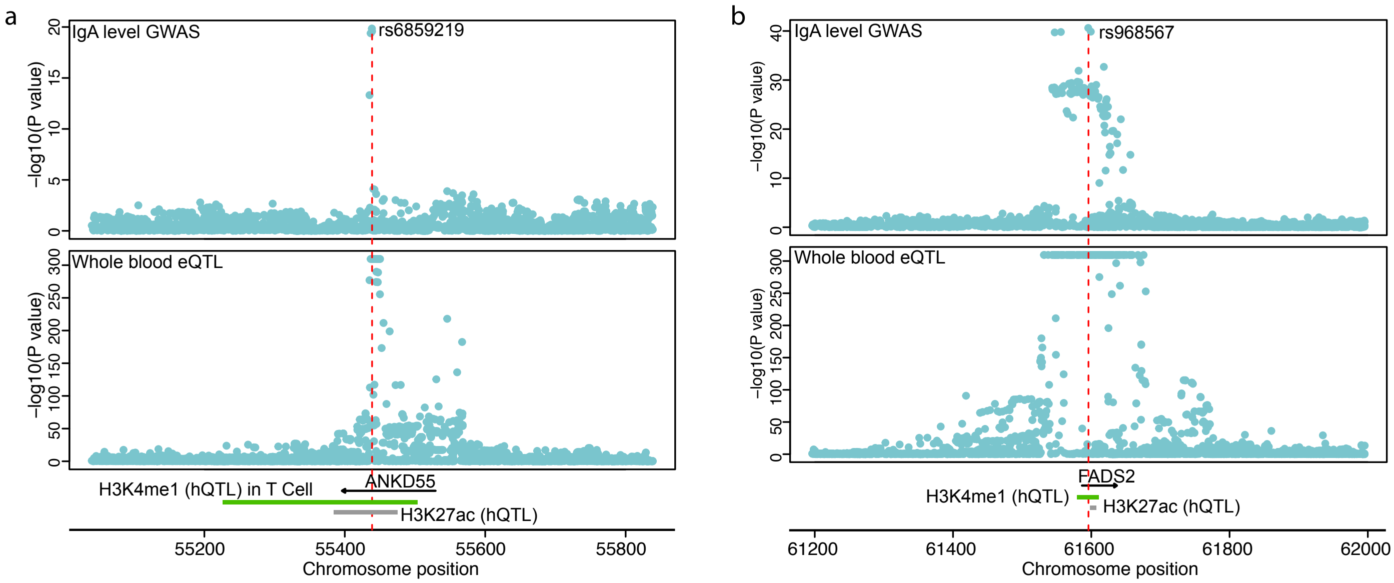

**Supplementary Figure 4. Integrative analysis of eQTL and hQTL for a) *ANKD55* and b) *FADS1/FDAS2* loci.** The upper and lower panels show the regional plots for IgA GWAS and eQTLs, respectively. The y-axis represents the -log10 of the p-value and x-axis shows the chromosome positions. The positions of candidate genes and hQTL peaks are depicted above the x-axis.

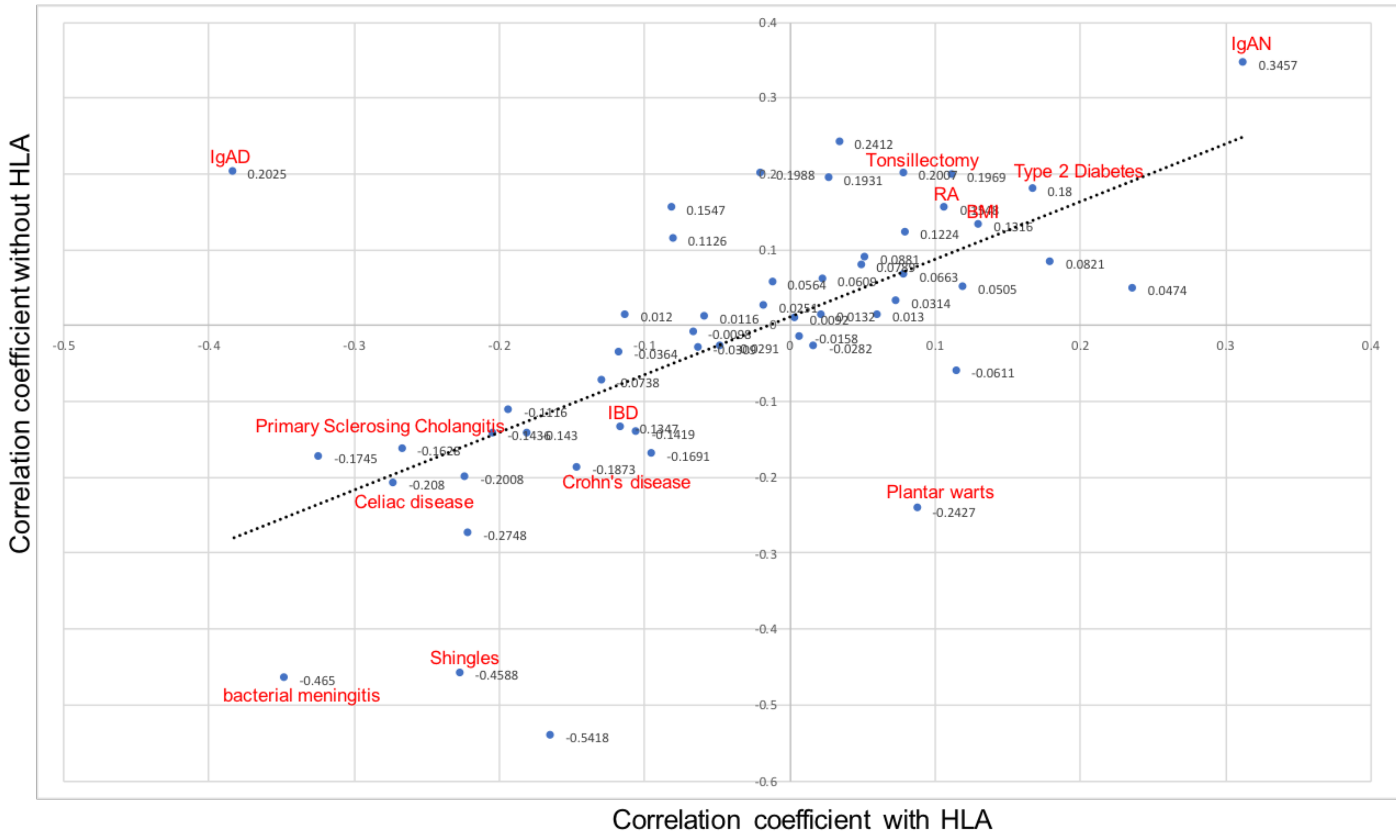

**Supplementary Figure 5. Genome-wide genetic correlations between serum IgA level and other complex traits with HLA region (x-axis) and without HLA region (y-axis).** Nominally significant correlations ($p<0.05$) are labeled in red. For two traits, IgA Deficiency (IgAD) and Plantar Warts, the inclusion of HLA region flips the sign of the genome-wide genetic correlation coefficient with serum IgA levels. IgAN: IgA nephropathy; IBD: Inflammatory Bowel Disease; RA: Rheumatoid Arthritis; BMI: Body Mass Index.

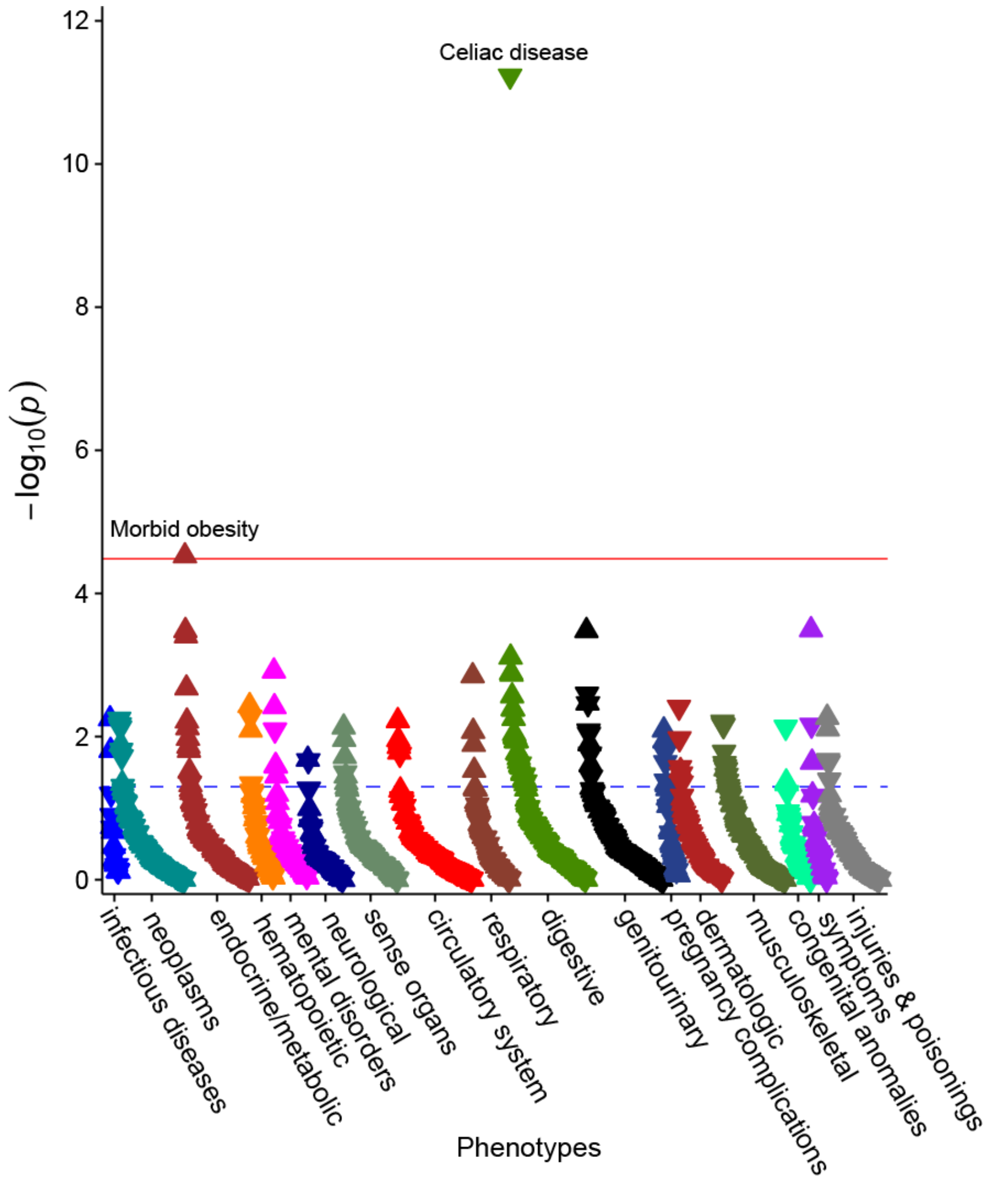

**Supplementary Figure 6. Meta-PheWAS of genome-wide polygenic score (GPS) for IgA levels without the HLA region across the UKBB and eMERGE-III biobanks (total N=556,656).** The y axis shows –log10 (P value) and the red line corresponds to the Bonferroni-corrected significance threshold for 1,523 phecodes (alpha=0.05/1,523=3.28x$10^{-5}$). Each triangle represents an association statistic for an individual phenotype (phecode) tested as an outcome against the GPS for IgA levels (without HLA) as a predictor; an upward triangle indicates a positive (risk) association, while a downward triangle indicates a negative (protective) association. All associations are adjusted for age, sex, site, genotyping batch, and principal components of ancestry as described in the Methods. The phenotypes are grouped by organ system (or relevant disease category) and sorted based on significance within each group.

### SUPPLEMENTAL TABLES

**Supplementary Table 1. Stepwise conditional analysis for genome-wide significant loci that exhibited more than one independently genome-wide significant risk variant.**

| Locus | CHR | BP (hg19) | SNP | Risk Allele | BETA | P-value | BETA (cond) | P-value (cond) |
|---|---|---|---|---|---|---|---|---|
| ***TNFSF4, TNFSF18*** | 1 | 173163568 | rs7518129 | G | 0.056 | 1.06E-16 | | |
| | 1 | 173172158 | rs4916314 | A | 0.061 | 3.46E-14 | 0.048 | 1.34E-09 |
| ***ELL2*** | 5 | 95277555 | rs3777175 | G | 0.084 | 7.81E-30 | | |
| | 5 | 95231115 | rs3777207 | T | 0.080 | 1.16E-25 | 0.080 | 6.44E-26 |
| | 5 | 95286609 | rs200990904 | C | 0.065 | 2.02E-14 | 0.020 | 3.00E-08 |
| ***HLA*** | 6 | 31106893 | rs1265094 | A | 0.076 | 1.86E-31 | | |
| | 6 | 32710119 | rs5018343 | A | 0.070 | 2.94E-24 | 0.076 | 4.96E-28 |
| | 6 | 32449523 | chr6:32449523 | T | 0.078 | 4.32E-25 | 0.078 | 3.60E-25 |
| | 6 | 29943715 | rs3823363 | C | 0.062 | 3.13E-22 | 0.059 | 8.42E-21 |
| | 6 | 31438243 | rs2596450 | G | 0.058 | 1.33E-17 | 0.058 | 3.41E-18 |
| | 6 | 32804299 | rs3819717 | C | 0.047 | 1.01E-13 | 0.052 | 2.24E-16 |
| | 6 | 31324152 | rs1050628 | G | 0.126 | 3.56E-16 | 0.123 | 1.45E-15 |
| | 6 | 32343369 | rs3117106 | T | 0.073 | 1.44E-12 | 0.075 | 1.69E-14 |
| ***FADS2, FADS1*** | 11 | 61595564 | rs968567 | T | 0.116 | 2.42E-41 | | |
| | 11 | 61557803 | rs102275 | C | 0.073 | 3.60E-29 | 0.060 | 1.60E-21 |
| ***HORMAD2, LIF*** | 22 | 30448399 | rs193473 | A | 0.067 | 1.58E-20 | | |
| | 22 | 30269907 | rs9614090 | A | 0.052 | 5.56E-16 | 0.036 | 3.65E-09 |

**Supplementary Table 2. Suggestive loci in the combined trans-ethnic meta-analysis (P<1.0E-06).**

| Locus | CHR | BP (hg19) | SNP | Risk Allele | BETA | P-value |
|---|---|---|---|---|---|---|
| ***TNFSF13*** | 17 | 7462969 | rs3803800 | A | 0.06 | 9.41E-08 |
| ***DLEU1*** | 13 | 50981003 | rs113429750 | C | 0.17 | 2.86E-07 |
| ***GUCY1A2*** | 11 | 106739921 | rs10431056 | C | 0.07 | 3.21E-07 |
| ***TNNT1*** | 19 | 55658222 | rs11550309 | T | 0.97 | 5.01E-07 |
| ***ARHGEF17*** | 11 | 73080563 | rs115997298 | G | 0.50 | 5.38E-07 |
| ***DZANK1, ZNF133*** | 20 | 18343798 | rs6045361 | G | 0.05 | 7.73E-07 |
| ***DCLK2*** | 4 | 150953126 | rs77297307 | C | 0.46 | 8.21E-07 |
| ***PDE10A*** | 6 | 166396027 | rs73260573 | G | 0.07 | 9.86E-07 |

**Supplementary Table 3. Pathway enrichment analysis**: top pathways enriched in genes encoded by the significant GWAS loci for serum IgA levels.

| Name | Source | P-value | FDR B&H | FDR B&Y | Bonferroni | No. of Genes (input) | No. of Genes (annotation) |
|---|---|---|---|---|---|---|---|
| Cytokine Signaling in Immune system | REACTOME | 1.17E-06 | 1.08E-03 | 7.99E-03 | 1.08E-03 | 24 | 763 |
| Cytokine-cytokine receptor interaction | KEGG | 5.23E-06 | 2.41E-03 | 1.79E-02 | 4.82E-03 | 13 | 270 |
| Signaling by Interleukins | REACTOME | 1.10E-05 | 3.18E-03 | 2.36E-02 | 1.01E-02 | 18 | 531 |
| TNFs bind their physiological receptors | REACTOME | 1.38E-05 | 3.18E-03 | 2.36E-02 | 1.27E-02 | 5 | 30 |
| IL-6-type cytokine receptor ligand interactions | REACTOME | 4.98E-05 | 9.19E-03 | 6.80E-02 | 4.59E-02 | 4 | 20 |
| Linoleic acid (LA) metabolism | REACTOME | 1.29E-04 | 1.99E-02 | 1.48E-01 | 1.20E-01 | 3 | 10 |
| Interleukin-6 family signaling | REACTOME | 1.70E-04 | 2.25E-02 | 1.66E-01 | 1.57E-01 | 4 | 27 |
| IL12-mediated signaling events | Pathway Interaction Database | 4.43E-04 | 3.96E-02 | 2.93E-01 | 4.09E-01 | 5 | 61 |
| alpha-linolenic acid (ALA) metabolism | REACTOME | 4.72E-04 | 3.96E-02 | 2.93E-01 | 4.36E-01 | 3 | 15 |
| TACI and BCMA stimulation of B cell immune responses. | MSigDB C2 BIOCARTA | 4.72E-04 | 3.96E-02 | 2.93E-01 | 4.36E-01 | 3 | 15 |

**Supplementary Table 4. Top ranked tissues and cell types based on DEPICT analysis.** The analysis was performed for 209 Medical Subject Heading (MeSH) tissue and cell type annotations, only results at nominal P<0.05 are displayed and sorted by the level of statistical significance, the results meeting FDR <5% highlighted in red (and graphically depicted in Figure 2b).

| MeSH term | MeSH first level term | MeSH second level term | MeSH first level term | Nominal P value | FDR < 5% |
|---|---|---|---|---|---|
| A15.378.316 | Bone Marrow Cells | Hemic and Immune Systems | Bone Marrow Cells | 4.89E-05 | Yes |
| A15.378 | Hematopoietic System | Hemic and Immune Systems | Hematopoietic System | 4.89E-05 | Yes |
| A15.145 | Blood | Hemic and Immune Systems | Blood | 2.77E-04 | Yes |
| A11.627 | Myeloid Cells | Cells | Myeloid Cells | 2.79E-04 | Yes |
| A15.382.680 | Phagocytes | Hemic and Immune Systems | Phagocytes | 3.31E-04 | Yes |
| A15.145.229 | Blood Cells | Hemic and Immune Systems | Blood Cells | 4.20E-04 | Yes |
| A15.145.300 | Fetal Blood | Hemic and Immune Systems | Fetal Blood | 6.36E-04 | Yes |
| A15.382.490.315.583 | Neutrophils | Hemic and Immune Systems | Neutrophils | 6.39E-04 | Yes |
| A11.118.637.415 | Granulocytes | Cells | Granulocytes | 6.57E-04 | Yes |
| A11.118.637 | Leukocytes | Cells | Leukocytes | 8.90E-04 | Yes |
| A02.835.583.443.800.800 | Synovial Fluid | Musculoskeletal System | Synovial Fluid | 0.0016 | Yes |
| A15.378.316.580 | Monocytes | Hemic and Immune Systems | Monocytes | 0.0019 | Yes |
| A15.382 | Immune System | Hemic and Immune Systems | Immune System | 0.003 | Yes |
| A11.066 | Antigen Presenting Cells | Cells | Antigen Presenting Cells | 0.003 | Yes |
| A15.382.812.260 | Dendritic Cells | Hemic and Immune Systems | Dendritic Cells | 0.003 | Yes |
| A15.382.520.604.700 | Spleen | Hemic and Immune Systems | Spleen | 0.004 | Yes |
| A15.382.812 | Mononuclear Phagocyte System | Hemic and Immune Systems | Mononuclear Phagocyte System | 0.005 | Yes |
| A15.382.520 | Lymphatic System | Hemic and Immune Systems | Lymphatic System | 0.008 | No |
| A10.549 | Lymphoid Tissue | Tissues | Lymphoid Tissue | 0.008 | No |
| A10.549.400 | Lymph Nodes | Tissues | Lymph Nodes | 0.009 | No |
| A02.835.232.043 | Bones of Lower Extremity | Musculoskeletal System | Bones of Lower Extremity | 0.010 | No |
| A15.145.229.637.555 | Leukocytes Mononuclear | Hemic and Immune Systems | Leukocytes Mononuclear | 0.012 | No |
| A11.118.637.555.567.569 | T Lymphocytes | Cells | T Lymphocytes | 0.012 | No |
| A02.835.232.043.300 | Foot Bones | Musculoskeletal System | Foot Bones | 0.013 | No |
| A02.835.232.043.300.710 | Tarsal Bones | Musculoskeletal System | Tarsal Bones | 0.013 | No |
| A11.118.637.555.567.569.200.700 | T Lymphocytes Regulatory | Cells | T Lymphocytes Regulatory | 0.016 | No |
| A09.371.060 | Anterior Eye Segment | Sense Organs | Anterior Eye Segment | 0.018 | No |
| A09.371.337.168 | Conjunctiva | Sense Organs | Conjunctiva | 0.018 | No |
| A09.371.337 | Eyelids | Sense Organs | Eyelids | 0.018 | No |
| A15.145.229.637.555.567.569.200 | CD4 Positive T Lymphocytes | Hemic and Immune Systems | CD4 Positive T Lymphocytes | 0.020 | No |
| A15.382.490.555.567.537 | Killer Cells Natural | Hemic and Immune Systems | Killer Cells Natural | 0.020 | No |
| A10.165 | Connective Tissue | Tissues | Connective Tissue | 0.020 | No |
| A15.382.216 | Bone Marrow | Hemic and Immune Systems | Bone Marrow | 0.021 | No |
| A02.835 | Skeleton | Musculoskeletal System | Skeleton | 0.024 | No |
| A02.835.232 | Bone and Bones | Musculoskeletal System | Bone and Bones | 0.024 | No |
| A15.382.812.522 | Macrophages | Hemic and Immune Systems | Macrophages | 0.029 | No |
| A11.872.378.294 | Lymphoid Progenitor Cells | Cells | Lymphoid Progenitor Cells | 0.032 | No |
| A11.118.637.555.567.562.440 | Precursor Cells B Lymphoid | Cells | Precursor Cells B Lymphoid | 0.032 | No |
| A04.531.520 | Nasal Mucosa | Respiratory System | Nasal Mucosa | 0.032 | No |
| A09.531 | Nose | Sense Organs | Nose | 0.032 | No |
| A10.615.550.760 | Respiratory Mucosa | Tissues | Respiratory Mucosa | 0.032 | No |
| A11.872.378 | Hematopoietic Stem Cells | Cells | Hematopoietic Stem Cells | 0.042 | No |

**Supplementary Table 5. Pleiotropy analysis.** We interrogated genome-wide significant GWAS loci for IgA levels against all previously studied GWAS phenotypes based on the GWAS catalogue; any reported trait associations for SNPs in LD ($R^2 > 0.5$) with the top variant at each GWAS locus are listed; the *SH2B3* locus exhibited the greatest range of pleiotropic associations.

| | Previously published GWAS associations | | | | | | | Associations with IgA levels | | LD with the top SNP |
|---|---|---|---|---|---|---|---|---|---|---|
| **Gene** | **Locus** | **Trait** | **PMID** | **Journal** | **Index SNP** | **P value** | **Annotation** | **Top SNP** | **P value** | **R2** |
| ANKRD55 | 5q11.2 | Rheumatoid arthritis | 20453842 | Nat Genet | rs6859219 | 1.00E-11 | intron | rs6859219 | 1.41E-20 | 1.00 |
| ANKRD55 | 5q11.2 | Multiple sclerosis | 27386562 | Sci Adv | rs6859219 | 8.00E-09 | intron | rs6859219 | 1.41E-20 | 1.00 |
| ANKRD55 | 5q11.2 | Crohn's disease | 23128233 | Nature | rs10065637 | 4.00E-12 | intron | rs6859219 | 1.41E-20 | 0.52 |
| BTG4 | 11q23.1 | Primary biliary cholangitis | 28062665 | Hum Mol Genet | rs4938534 | 1.00E-08 | intron | rs4938518 | 7.01E-16 | 0.84 |
| CTF1 | 16p11.2 | Diastolic blood pressure | 27841878 | Nat Genet | rs72799341 | 4.00E-09 | intron | rs1458201 | 2.02E-11 | 0.94 |
| ELL2 | 5q15 | Serum total protein level | 23022100 | Am J Hum Genet | rs3777200 | 1.00E-08 | intron | rs3777175 | 7.81E-30 | 0.74 |
| ELL2 | 5q15 | Albumin-globulin ratio | 29403010 | Nat Genet | rs3777184 | 8.00E-14 | intron | rs3777175 | 7.81E-30 | 0.72 |
| ELL2 | 5q15 | Multiple myeloma | 27363682 | Nat Commun | rs1423269 | 2.00E-11 | intron | rs3777175 | 7.81E-30 | 0.69 |
| ELL2 | 5q15 | Multiple myeloma and monoclonal gammopathy | 26007630 | Nat Commun | rs56219066 | 2.00E-10 | intron | rs3777175 | 7.81E-30 | 0.67 |
| ELL2 | 5q15 | Mean corpuscular hemoglobin | 30595370 | Am J Hum Genet | rs3756703 | 1.00E-09 | intron | rs3777175 | 7.81E-30 | 0.61 |
| FADS2 | 11q12.2 | Fatty acid desaturase activity (serum) | 30453627 | Nutrients | rs968567 | 5.00E-49 | 5_prime_UTR | rs968567 | 2.42E-41 | 1.00 |
| FADS2 | 11q12.2 | Blood metabolite levels | 24816252 | Nat Genet | rs968567 | 2.00E-21 | 5_prime_UTR | rs968567 | 2.42E-41 | 1.00 |
| FADS2 | 11q12.2 | Glycerophospholipid levels | 26068415 | Nat Commun | rs968567 | 1.00E-18 | 5_prime_UTR | rs968567 | 2.42E-41 | 1.00 |
| FADS2 | 11q12.2 | Serum metabolite ratios in chronic kidney disease | 29545352 | J Am Soc Nephrol | rs968567 | 5.00E-14 | 5_prime_UTR | rs968567 | 2.42E-41 | 1.00 |
| FADS2 | 11q12.2 | Rheumatoid arthritis | 24390342 | Nature | rs968567 | 2.00E-08 | 5_prime_UTR | rs968567 | 2.42E-41 | 1.00 |
| FADS2 | 11q12.2 | Hair color | 30595370 | Am J Hum Genet | rs61896141 | 6.00E-21 | intron | rs968567 | 2.42E-41 | 0.98 |
| FADS2 | 11q12.2 | Blond vs. brown/black hair color | 30531825 | Nat Commun | rs61896141 | 1.00E-18 | intron | rs968567 | 2.42E-41 | 0.98 |
| FADS2 | 11q12.2 | Serum metabolite ratios in chronic kidney disease | 29545352 | J Am Soc Nephrol | rs7943728 | 4.00E-19 | intron | rs968567 | 2.42E-41 | 0.97 |
| FADS2 | 11q12.2 | Sum basophil neutrophil counts | 27863252 | Cell | rs61897795 | 4.00E-12 | intron | rs968567 | 2.42E-41 | 0.83 |
| FADS2 | 11q12.2 | Neutrophil count | 27863252 | Cell | rs61897795 | 6.00E-12 | intron | rs968567 | 2.42E-41 | 0.83 |
| HDAC7 | 12q13.11 | Platelet count | 27863252 | Cell | rs73109811 | 6.00E-18 | intron | rs7487637 | 9.97E-15 | 0.65 |
| HDAC7 | 12q13.11 | Chronic obstructive pulmonary disease or high blood pressure | 30940143 | Respir Res | rs11168245 | 7.00E-11 | intron | rs7487637 | 9.97E-15 | 0.55 |
| HDAC7 | 12q13.11 | Systolic blood pressure | 27841878 | Nat Genet | rs11168244 | 2.00E-10 | intron | rs7487637 | 9.97E-15 | 0.55 |
| HDAC7 | 12q13.11 | Diastolic blood pressure | 27841878 | Nat Genet | rs11168244 | 5.00E-08 | intron | rs7487637 | 9.97E-15 | 0.55 |
| HDAC7 | 12q13.11 | Allergic disease (asthma, hay fever or eczema) | 29083406 | Nat Genet | rs55726902 | 3.00E-16 | intron | rs7487637 | 9.97E-15 | 0.53 |
| HDAC7 | 12q13.11 | Varicose veins | 30566020 | Circulation | rs73107980 | 8.00E-11 | intron | rs7487637 | 9.97E-15 | 0.52 |
| HORMAD2 | 22q12.2 | Tonsillectomy | 28928442 | Nat Commun | rs9625935 | 6.00E-34 | intron | rs193473 | 1.58E-20 | 0.79 |
| HORMAD2 | 22q12.2 | Lung cancer | 21725308 | Nat Genet | rs36600 | 6.00E-13 | intron | rs193473 | 1.58E-20 | 0.76 |
| HORMAD2 | 22q12.2 | Chronic inflammatory diseases | 26974007 | Nat Genet | rs140135 | 6.00E-09 | intron | rs193473 | 1.58E-20 | 0.63 |
| OVOL1 | 11q13.1 | Blood protein levels | 29875488 | Nature | rs10896045 | 1.00E-11 | intron | rs10896045 | 2.57E-22 | 1.00 |
| OVOL1 | 11q13.1 | Eczema | 30595370 | Am J Hum Genet | rs10791824 | 8.00E-24 | intron | rs10896045 | 2.57E-22 | 0.68 |
| OVOL1 | 11q13.1 | Atopic dermatitis | 26482879 | Nat Genet | rs10791824 | 2.00E-19 | intron | rs10896045 | 2.57E-22 | 0.68 |
| RUNX3 | 1p36.11 | IgG glycosylation patterns | 29535710 | Front Immunol | rs16830188 | 4.00E-10 | intergenic | rs188468174 | 3.42E-92 | 1.00 |
| TNFSF4 | 1q25.1 | Eczema | 30595370 | Am J Hum Genet | rs7518129 | 4.00E-18 | intron | rs7518129 | 1.06E-16 | 1.00 |
| TNFSF4 | 1q25.1 | Asthma | 27182965 | Nat Genet | rs6691738 | 3.00E-08 | intergenic | rs7518129 | 1.06E-16 | 0.87 |
| TNFSF4 | 1q25.1 | Narcolepsy | 23459209 | PLoS Genet | rs7553711 | 4.00E-08 | regulatory | rs7518129 | 1.06E-16 | 0.87 |
| TRAF3 | 14q32.32 | Eczema | 30595370 | Am J Hum Genet | rs12886625 | 9.00E-13 | intron | rs12147883 | 5.42E-14 | 0.91 |
| TRAF3 | 14q32.32 | Albumin-globulin ratio | 29403010 | Nat Genet | rs71421264 | 3.00E-19 | intron | rs12147883 | 5.42E-14 | 0.82 |
| TRAF3 | 14q32.32 | Immunoglobulin measurement (zinc sulfate turbidity test) | 29403010 | Nat Genet | rs1051750 | 1.00E-09 | 5_prime_UTR | rs12147883 | 5.42E-14 | 0.82 |
| TRAF3 | 14q32.32 | Body mass index | 30595370 | Am J Hum Genet | rs3803286 | 3.00E-21 | intron | rs12147883 | 5.42E-14 | 0.72 |
| TRAF3 | 14q32.32 | Non-albumin protein levels | 29403010 | Nat Genet | rs12432676 | 2.00E-24 | intron | rs12147883 | 5.42E-14 | 0.68 |
| TRAF3 | 14q32.32 | Self-reported math ability (MTAG) | 30038396 | Nat Genet | rs12434396 | 3.00E-08 | intron | rs12147883 | 5.42E-14 | 0.68 |
| TRAF3 | 14q32.32 | Serum total protein level | 29403010 | Nat Genet | rs8022180 | 3.00E-20 | intron | rs12147883 | 5.42E-14 | 0.66 |
| TRAF3 | 14q32.32 | Multiple sclerosis | 24076602 | Nat Genet | rs12148050 | 5.00E-13 | intron | rs12147883 | 5.42E-14 | 0.66 |
| SH2B3 | 12q24.12 | Beta-2 microglobulin plasma levels | 23417110 | Hum Genet | rs3184504 | 3.00E-08 | missense | rs10774624 | 1.37E-13 | 0.94 |
| SH2B3 | 12q24.12 | Blood metabolite levels | 24816252 | Nat Genet | rs3184504 | 6.00E-18 | missense | rs10774624 | 1.37E-13 | 0.94 |
| SH2B3 | 12q24.12 | Blood pressure | 21909110 | Nat Genet | rs653178 | 7.00E-20 | intron | rs10774624 | 1.37E-13 | 0.93 |
| SH2B3 | 12q24.12 | Blood protein levels | 29875488 | Nature | rs3184504 | 2.00E-14 | missense | rs10774624 | 1.37E-13 | 0.94 |
| SH2B3 | 12q24.12 | Body mass index | 30108127 | Genetics | rs3184504 | 6.00E-09 | missense | rs10774624 | 1.37E-13 | 0.94 |
| SH2B3 | 12q24.12 | Cardiovascular disease | 30595370 | Am J Hum Genet | rs3184504 | 3.00E-34 | missense | rs10774624 | 1.37E-13 | 0.94 |
| SH2B3 | 12q24.12 | Celiac disease | 22057235 | Nat Genet | rs3184504 | 5.00E-21 | missense | rs10774624 | 1.37E-13 | 0.94 |
| SH2B3 | 12q24.12 | Celiac disease or Rheumatoid arthritis | 21383967 | PLoS Genet | rs653178 | 3.00E-19 | intron | rs10774624 | 1.37E-13 | 0.93 |
| SH2B3 | 12q24.12 | Cholesterol, total | 20686565 | Nature | rs11065987 | 7.00E-12 | intergenic | rs10774624 | 1.37E-13 | 0.85 |
| SH2B3 | 12q24.12 | Chronic inflammatory diseases | 26974007 | Nat Genet | rs3184504 | 4.00E-10 | missense | rs10774624 | 1.37E-13 | 0.94 |
| SH2B3 | 12q24.12 | Chronic kidney disease | 20383146 | Nat Genet | rs653178 | 4.00E-11 | intron | rs10774624 | 1.37E-13 | 0.93 |
| SH2B3 | 12q24.12 | Colorectal cancer | 29917119 | J Natl Cancer Inst | rs3184504 | 2.00E-10 | missense | rs10774624 | 1.37E-13 | 0.94 |
| SH2B3 | 12q24.12 | Colorectal or endometrial cancer | 26621817 | Sci Rep | rs3184504 | 8.00E-09 | missense | rs10774624 | 1.37E-13 | 0.94 |
| SH2B3 | 12q24.12 | Coronary artery disease | 29212778 | Circ Res | rs3184504 | 5.00E-30 | missense | rs10774624 | 1.37E-13 | 0.94 |
| SH2B3 | 12q24.12 | Diastolic blood pressure | 27618452 | Nat Genet | rs3184504 | 1.00E-21 | missense | rs10774624 | 1.37E-13 | 0.94 |
| SH2B3 | 12q24.12 | Eczema | 30595370 | Am J Hum Genet | rs653178 | 4.00E-19 | intron | rs10774624 | 1.37E-13 | 0.93 |
| SH2B3 | 12q24.12 | Endometrial cancer | 30093612 | Nat Commun | rs3184504 | 1.00E-10 | missense | rs10774624 | 1.37E-13 | 0.94 |
| SH2B3 | 12q24.12 | Eosinophil counts | 30595370 | Am J Hum Genet | rs10774624 | 1.00E-300 | intron | rs10774624 | 1.37E-13 | 1.00 |
| SH2B3 | 12q24.12 | Eosinophil percentage of granulocytes | 27863252 | Cell | rs653178 | 4.00E-126 | intron | rs10774624 | 1.37E-13 | 0.93 |
| SH2B3 | 12q24.12 | Eosinophil percentage of white cells | 27863252 | Cell | rs653178 | 3.00E-120 | intron | rs10774624 | 1.37E-13 | 0.93 |
| SH2B3 | 12q24.12 | Fibrinogen levels | 26561523 | Hum Mol Genet | rs7310615 | 2.00E-13 | intron | rs10774624 | 1.37E-13 | 0.94 |
| SH2B3 | 12q24.12 | Glaucoma (primary open-angle) | 26752265 | Nat Genet | rs7137828 | 9.00E-10 | intron | rs10774624 | 1.37E-13 | 0.94 |
| SH2B3 | 12q24.12 | Glycated hemoglobin levels | 28898252 | PLoS Med | rs10774625 | 1.00E-08 | intron | rs10774624 | 1.37E-13 | 0.94 |
| SH2B3 | 12q24.12 | Granulocyte count | 27863252 | Cell | rs3184504 | 3.00E-21 | missense | rs10774624 | 1.37E-13 | 0.94 |
| SH2B3 | 12q24.12 | HDL cholesterol | 30275531 | Nat Genet | rs4766578 | 1.00E-14 | intron | rs10774624 | 1.37E-13 | 0.94 |
| SH2B3 | 12q24.12 | Hematocrit | 27863252 | Cell | rs3184504 | 8.00E-72 | missense | rs10774624 | 1.37E-13 | 0.94 |
| SH2B3 | 12q24.12 | Hemoglobin concentration | 27863252 | Cell | rs3184504 | 1.00E-74 | missense | rs10774624 | 1.37E-13 | 0.94 |
| SH2B3 | 12q24.12 | Hemoglobin levels | 28017375 | Am J Hum Genet | rs3184504 | 1.00E-10 | missense | rs10774624 | 1.37E-13 | 0.94 |
| SH2B3 | 12q24.12 | High density lipoprotein cholesterol levels | 29507422 | Nat Genet | rs3184504 | 9.00E-12 | missense | rs10774624 | 1.37E-13 | 0.94 |
| SH2B3 | 12q24.12 | High light scatter reticulocyte count | 27863252 | Cell | rs4766578 | 3.00E-75 | intron | rs10774624 | 1.37E-13 | 0.94 |
| SH2B3 | 12q24.12 | High light scatter reticulocyte percentage of red cells | 27863252 | Cell | rs4766578 | 2.00E-60 | intron | rs10774624 | 1.37E-13 | 0.94 |
| SH2B3 | 12q24.12 | Hypothyroidism | 27182965 | Nat Genet | rs10774625 | 1.00E-26 | intron | rs10774624 | 1.37E-13 | 0.94 |
| SH2B3 | 12q24.12 | Immature fraction of reticulocytes | 27863252 | Cell | rs4766578 | 4.00E-19 | intron | rs10774624 | 1.37E-13 | 0.94 |
| SH2B3 | 12q24.12 | Inflammatory bowel disease | 28067908 | Nat Genet | rs653178 | 2.00E-09 | intron | rs10774624 | 1.37E-13 | 0.93 |
| SH2B3 | 12q24.12 | Intraocular pressure | 30054594 | Nat Genet | rs11065979 | 3.00E-09 | intergenic | rs10774624 | 1.37E-13 | 0.88 |
| SH2B3 | 12q24.12 | Intraocular pressure | 29785010 | Nat Genet | rs10774624 | 3.00E-10 | intron | rs10774624 | 1.37E-13 | 1.00 |
| SH2B3 | 12q24.12 | Juvenile idiopathic arthritis | 23603761 | Nat Genet | rs7137828 | 2.00E-09 | intron | rs10774624 | 1.37E-13 | 0.94 |
| SH2B3 | 12q24.12 | Latent autoimmune diabetes vs. type 2 diabetes | 30254083 | Diabetes Care | rs3184504 | 2.00E-08 | missense | rs10774624 | 1.37E-13 | 0.94 |
| SH2B3 | 12q24.12 | LDL cholesterol | 30275531 | Nat Genet | rs653178 | 4.00E-20 | intron | rs10774624 | 1.37E-13 | 0.93 |
| SH2B3 | 12q24.12 | Left ventricle diastolic internal dimension | 28394258 | J Clin Invest | rs10774625 | 1.00E-08 | intron | rs10774624 | 1.37E-13 | 0.94 |
| SH2B3 | 12q24.12 | Low density lipoprotein cholesterol levels | 29507422 | Nat Genet | rs3184504 | 2.00E-12 | missense | rs10774624 | 1.37E-13 | 0.94 |

| | | | | | | | | | | |
|---|---|---|---|---|---|---|---|---|---|---|
| SH2B3 | 12q24.12 | Lymphocyte counts | 27863252 | Cell | rs3184504 | 7.00E-134 | missense | rs10774624 | 1.37E-13 | 0.94 |
| SH2B3 | 12q24.12 | Mean arterial pressure | 27618448 | Nat Genet | rs653178 | 1.00E-23 | intron | rs10774624 | 1.37E-13 | 0.93 |
| SH2B3 | 12q24.12 | Mean corpuscular hemoglobin | 30595370 | Am J Hum Genet | rs597808 | 4.00E-24 | intron | rs10774624 | 1.37E-13 | 0.93 |
| SH2B3 | 12q24.12 | Monocyte count | 27863252 | Cell | rs653178 | 1.00E-43 | intron | rs10774624 | 1.37E-13 | 0.93 |
| SH2B3 | 12q24.12 | Myeloid white cell count | 27863252 | Cell | rs3184504 | 1.00E-26 | missense | rs10774624 | 1.37E-13 | 0.94 |
| SH2B3 | 12q24.12 | Myocardial infarction | 26343387 | Nat Genet | rs653178 | 3.00E-11 | intron | rs10774624 | 1.37E-13 | 0.93 |
| SH2B3 | 12q24.12 | Neutrophil count | 27863252 | Cell | rs3184504 | 4.00E-12 | missense | rs10774624 | 1.37E-13 | 0.94 |
| SH2B3 | 12q24.12 | Neutrophil percentage of granulocytes | 27863252 | Cell | rs653178 | 5.00E-108 | intron | rs10774624 | 1.37E-13 | 0.93 |
| SH2B3 | 12q24.12 | Parental longevity (combined parental age at death) | 29227965 | Aging (Albany NY) | rs7137828 | 3.00E-08 | intron | rs10774624 | 1.37E-13 | 0.94 |
| SH2B3 | 12q24.12 | Parental longevity (combined parental attained age) | 29227965 | Aging (Albany NY) | rs7137828 | 3.00E-14 | intron | rs10774624 | 1.37E-13 | 0.94 |
| SH2B3 | 12q24.12 | Parental longevity (combined parental attained age) | 29227965 | Aging (Albany NY) | rs11065979 | 2.00E-12 | intergenic | rs10774624 | 1.37E-13 | 0.88 |
| SH2B3 | 12q24.12 | Parental longevity (father's age at death) | 29227965 | Aging (Albany NY) | rs3184504 | 4.00E-08 | missense | rs10774624 | 1.37E-13 | 0.94 |
| SH2B3 | 12q24.12 | Parental longevity (father's attained age) | 29227965 | Aging (Albany NY) | rs7137828 | 3.00E-12 | intron | rs10774624 | 1.37E-13 | 0.94 |
| SH2B3 | 12q24.12 | Platelet count | 27863252 | Cell | rs3184504 | 6.00E-180 | missense | rs10774624 | 1.37E-13 | 0.94 |
| SH2B3 | 12q24.12 | Plateletcrit | 27863252 | Cell | rs3184504 | 5.00E-216 | missense | rs10774624 | 1.37E-13 | 0.94 |
| SH2B3 | 12q24.12 | Primary biliary cholangitis | 26394269 | Nat Commun | rs11065987 | 3.00E-08 | intergenic | rs10774624 | 1.37E-13 | 0.85 |
| SH2B3 | 12q24.12 | Primary biliary cirrhosis | 22961000 | Nat Genet | rs11065979 | 3.00E-09 | intergenic | rs10774624 | 1.37E-13 | 0.88 |
| SH2B3 | 12q24.12 | Primary sclerosing cholangitis | 23603763 | Nat Genet | rs3184504 | 6.00E-11 | missense | rs10774624 | 1.37E-13 | 0.94 |
| SH2B3 | 12q24.12 | Psoriasis | 28537254 | Nat Commun | rs11065979 | 2.00E-08 | intergenic | rs10774624 | 1.37E-13 | 0.88 |
| SH2B3 | 12q24.12 | Reaction time | 29844566 | Nat Commun | rs4766578 | 1.00E-08 | intron | rs10774624 | 1.37E-13 | 0.94 |
| SH2B3 | 12q24.12 | Red blood cell count | 27863252 | Cell | rs3184504 | 2.00E-43 | missense | rs10774624 | 1.37E-13 | 0.94 |
| SH2B3 | 12q24.12 | Red blood cell traits | 23222517 | Nature | rs3184504 | 4.00E-19 | missense | rs10774624 | 1.37E-13 | 0.94 |
| SH2B3 | 12q24.12 | Reticulocyte count | 27863252 | Cell | rs3184504 | 7.00E-91 | missense | rs10774624 | 1.37E-13 | 0.94 |
| SH2B3 | 12q24.12 | Reticulocyte fraction of red cells | 27863252 | Cell | rs4766578 | 2.00E-69 | intron | rs10774624 | 1.37E-13 | 0.94 |
| SH2B3 | 12q24.12 | Retinal vascular caliber | 21060863 | PLoS Genet | rs10774625 | 2.00E-13 | intron | rs10774624 | 1.37E-13 | 0.94 |
| SH2B3 | 12q24.12 | Rheumatoid arthritis | 24390342 | Nature | rs10774624 | 7.00E-09 | intron | rs10774624 | 1.37E-13 | 1.00 |
| SH2B3 | 12q24.12 | Sarcoidosis | 26051272 | Am J Resp Crit Care | rs653178 | 2.00E-10 | intron | rs10774624 | 1.37E-13 | 0.93 |
| SH2B3 | 12q24.12 | Smoking status (ever vs never smokers) | 30643258 | Nat Genet | rs597808 | 2.00E-11 | intron | rs10774624 | 1.37E-13 | 0.93 |
| SH2B3 | 12q24.12 | Stroke | 29531354 | Nat Genet | rs3184504 | 9.00E-12 | missense | rs10774624 | 1.37E-13 | 0.94 |
| SH2B3 | 12q24.12 | Sum basophil neutrophil counts | 27863252 | Cell | rs3184504 | 4.00E-13 | missense | rs10774624 | 1.37E-13 | 0.94 |
| SH2B3 | 12q24.12 | Sum eosinophil basophil counts | 27863252 | Cell | rs653178 | 7.00E-167 | intron | rs10774624 | 1.37E-13 | 0.93 |
| SH2B3 | 12q24.12 | Sum neutrophil eosinophil counts | 27863252 | Cell | rs3184504 | 3.00E-20 | missense | rs10774624 | 1.37E-13 | 0.94 |
| SH2B3 | 12q24.12 | Systemic lupus erythematosus | 28714469 | Nat Commun | rs653178 | 7.00E-09 | intron | rs10774624 | 1.37E-13 | 0.93 |
| SH2B3 | 12q24.12 | Systolic blood pressure | 30578418 | Nat Genet | rs10774624 | 3.00E-19 | intron | rs10774624 | 1.37E-13 | 1.00 |
| SH2B3 | 12q24.12 | Tetralogy of Fallot | 23297363 | Hum Mol Genet | rs11065987 | 8.00E-11 | intergenic | rs10774624 | 1.37E-13 | 0.85 |
| SH2B3 | 12q24.12 | Thyroid peroxidase antibody positivity | 24586183 | PLoS Genet | rs653178 | 1.00E-09 | intron | rs10774624 | 1.37E-13 | 0.93 |
| SH2B3 | 12q24.12 | Tonsillectomy | 27182965 | Nat Genet | rs3184504 | 3.00E-10 | missense | rs10774624 | 1.37E-13 | 0.94 |
| SH2B3 | 12q24.12 | Tonsillectomy | 28928442 | Nat Commun | rs3184504 | 3.00E-10 | missense | rs10774624 | 1.37E-13 | 0.94 |
| SH2B3 | 12q24.12 | Total cholesterol levels | 30275531 | Nat Genet | rs653178 | 2.00E-22 | intron | rs10774624 | 1.37E-13 | 0.93 |
| SH2B3 | 12q24.12 | Type 1 diabetes | 25751624 | Nat Genet | rs653178 | 2.00E-44 | intron | rs10774624 | 1.37E-13 | 0.93 |
| SH2B3 | 12q24.12 | Urate levels | 23263486 | Nat Genet | rs653178 | 7.00E-12 | intron | rs10774624 | 1.37E-13 | 0.93 |
| SH2B3 | 12q24.12 | Vitiligo | 27723757 | Nat Genet | rs10774624 | 6.00E-23 | intron | rs10774624 | 1.37E-13 | 1.00 |
| SH2B3 | 12q24.12 | Waist-hip ratio | 30595370 | Am J Hum Genet | rs597808 | 3.00E-09 | intron | rs10774624 | 1.37E-13 | 0.93 |
| SH2B3 | 12q24.12 | White blood cell count | 27863252 | Cell | rs3184504 | 9.00E-70 | missense | rs10774624 | 1.37E-13 | 0.94 |

**Supplementary Table 6. ANNOVAR annotation of the top significant SNPs and their proxies (R2>0.5).** * indicates top SNP in GWAS

| Locus | SNP | Chr. | Position (hg19) | Risk allele | Non-risk allele | Beta | P (discovery) | $R^2$ with top SNP | Annotation |
|---|---|---|---|---|---|---|---|---|---|
| OVOL1 | rs10896045 | 11 | 65555524 | **a** | g | 0.066 | 2.57E-22 | 1.00* | intronic |
| POU2AF1 | rs4938518 | 11 | 111267394 | t | c | 0.056 | 7.01E-16 | 1.00* | intergenic |
| HDAC7 | rs7487637 | 12 | 48214825 | a | g | -0.054 | 9.97E-15 | 1.00* | intergenic |
| RCOR1,TRAF3 | rs12147883 | 14 | 103239630 | t | c | -0.048 | 5.42E-14 | 1.00* | intergenic b/w TRAF3 and RCOR1 |
| RCOR1,TRAF3 | rs1051750 | 14 | 103243946 | t | c | -0.045 | 4.72E-11 | 0.82 | UTR5 of TRAF3 |
| RUNX2 | rs1200427 | 6 | 45526470 | a | t | -0.059 | 6.85E-14 | 1.00* | intergenic b/w RUNX2 and CLIC5 |
| RUNX2 | rs1200428 | 6 | 45518202 | t | g | 0.035 | 8.47E-03 | 0.51 | UTR3 of RUNX2 |
| ZP3,SSC4D | rs55722505 | 7 | 76034150 | c | g | 0.048 | 8.61E-14 | 1.00* | intronic |
| ZP3,SSC4D | rs10261314 | 7 | 76023029 | t | c | 0.044 | 3.95E-12 | 0.62 | synonymous in SSC4D |
| LITAF | rs113962704 | 16 | 11717832 | a | t | -0.054 | 1.91E-12 | 1.00* | intergenic b/w LITAF and SNN |
| CTF1 | rs1458201 | 16 | 30916129 | a | g | 0.052 | 2.02E-11 | 1.00* | intergenic b/w CTF1 and FBXL19-AS1 |
| CTF1 | rs35675346 | 16 | 30936081 | a | g | 0.028 | 0.02305 | 0.57 | missense in FBXL19 |
| FILNC1 | rs17069163 | 6 | 139975943 | t | c | 0.050 | 5.94E-11 | 1.00* | intergenic b/w LINC01625 and LOC100132735 |
| IL1R1 | rs13427957 | 2 | 102689031 | t | c | -0.040 | 6.19E-10 | 1.00* | intronic |
| RUNX3 | rs188468174 | 1 | 25291697 | t | c | -0.876 | 3.42E-92 | 1.00* | intergenic b/w RUNX3 and MIR4425 |
| FADS2 | rs968567 | 11 | 61595564 | t | c | 0.116 | 2.42E-41 | 1.00* | UTR5 of FADS2 |
| ELL2 | rs3777175 | 5 | 95277555 | a | g | -0.084 | 7.81E-30 | 1.00* | intergenic |
| ELL2 | rs3777203 | 5 | 95234377 | t | c | 0.076 | 9.20E-28 | 0.74 | synonymous in ELL2 |
| ELL2 | rs17085249 | 5 | 95236415 | a | g | 0.076 | 8.59E-28 | 0.70 | synonymous in ELL2 |
| ELL2 | rs3815768 | 5 | 95236459 | t | c | 0.076 | 2.91E-27 | 0.69 | missense in ELL2 (BENIGN, 0.0) |
| ELL2 | rs3777202 | 5 | 95234392 | a | c | -0.076 | 5.22E-28 | 0.67 | synonymous in ELL2 |
| ELL2 | rs17085231 | 5 | 95222156 | t | c | 0.077 | 4.94E-28 | 0.65 | UTR3 of ELL2 |
| ELL2 | rs11135442 | 5 | 95222511 | t | c | -0.077 | 1.31E-28 | 0.60 | UTR3 of ELL2 |
| ELL2 | rs3777204 | 5 | 95234350 | a | g | -0.074 | 1.23E-26 | 0.56 | synonymous in ELL2 |
| ELL2 | rs1043381 | 5 | 95221337 | t | c | 0.073 | 5.44E-26 | 0.50 | UTR3 of ELL2 |
| ANKRD55 | rs6859219 | 5 | 55438580 | a | c | -0.073 | 1.41E-20 | 1.00* | intronic |
| HORMAD2 | rs193473 | 22 | 30448399 | a | g | 0.067 | 1.58E-20 | 1.00* | intronic |
| TNFSF8 | rs3181356 | 9 | 117692882 | t | c | 0.069 | 1.13E-18 | 1.00 | upstream TNFSF8 (dist=7) |
| TNFSF8 | rs55918341 | 9 | 117547198 | ca | c | -0.026 | 0.3385 | 0.68 | UTR3 of TNFSF15 |
| TNFSF8 | rs7853287 | 9 | 117549327 | a | g | 0.078 | 3.95E-09 | 0.68 | UTR3 of TNFSF15 |
| TNFSF4 | rs7518129 | 1 | 173163568 | a | g | -0.056 | 1.06E-16 | 1.00* | intergenic |
| TNFSF4 | rs7514229 | 1 | 173154304 | t | g | 0.055 | 6.13E-16 | 0.83 | UTR3 of TNFSF4 |
| SH2B3 | rs10774624 | 12 | 111833788 | a | g | -0.050 | 1.37E-13 | 1.00* | intergenic b/w FAM109A and SH2B3 |
| SH2B3 | rs3184504 | 12 | 111884608 | t | c | 0.048 | 6.07E-13 | 0.94 | missense in SH2B3 (BENIGN, 0.0) |

**Supplementary Table 7. Colocalization analysis with eQTLs from whole blood and 13 primary immune cell types.** Posterior probabilities of sharing and not sharing the same causal variant between GWAS and eQTL are designated as PP4 and PP3, respectively. The designation of a "concordant direction" indicates that IgA level increasing allele was also associated with higher expression of corresponding genes; "opposite direction" indicates that the IgA increasing allele was associated with lower expression of corresponding genes.

| Cell type | Chr | GWAS Locus Region (Start-End) | GWAS Lead SNP | GWAS p Value | eSNP | eSNP p Value | PP3 | PP4 | Gene | Direction |
|---|---|---|---|---|---|---|---|---|---|---|
| B cell NAÏVE | 6 | 30706942-31506854 | rs1265094 | 1.86E-31 | rs720467 | 2.95E-07 | 0.34 | 0.56 | TCF19 | Opposite |
| | 7 | 75634687-76433936 | rs55722505 | 8.61E-14 | rs201874621 | 9.30E-09 | 0.34 | 0.58 | POMZP3 | Concordant |
| | 11 | 61196289-61995513 | rs968567 | 2.42E-41 | rs174528 | 1.70E-11 | 0.16 | 0.84 | FADS1 | Concordant |
| | 11 | 61196289-61995513 | rs968567 | 2.42E-41 | rs7943728 | 2.18E-20 | 0.00 | 1.00 | FADS2 | Concordant |
| CD4 NAÏVE | 11 | 61196289-61995513 | rs968567 | 2.42E-41 | rs174576 | 5.65E-18 | 0.23 | 0.77 | FADS1 | Concordant |
| | 11 | 61196289-61995513 | rs968567 | 2.42E-41 | rs7943728 | 7.33E-16 | 0.00 | 1.00 | FADS2 | Concordant |
| | 14 | 102840084-103639456 | rs12147883 | 5.42E-14 | rs4906263 | 9.77E-05 | 0.07 | 0.61 | TRAF3 | Concordant |
| CD4 STIM | 11 | 61196289-61995513 | rs968567 | 2.42E-41 | rs7943728 | 5.22E-13 | 0.01 | 0.99 | FADS2 | Concordant |
| CD8 NAÏVE | 6 | 30706942-31506854 | rs1265094 | 1.86E-31 | rs2535324 | 1.84E-04 | 0.01 | 0.55 | HCG4 | Concordant |
| | 7 | 75634687-76433936 | rs55722505 | 8.61E-14 | rs201874621 | 3.86E-11 | 0.41 | 0.58 | POMZP3 | Concordant |
| | 11 | 61196289-61995513 | rs968567 | 2.42E-41 | rs7943728 | 5.61E-16 | 0.02 | 0.98 | FADS1 | Concordant |
| | 11 | 61196289-61995513 | rs968567 | 2.42E-41 | rs7943728 | 9.88E-16 | 0.00 | 1.00 | FADS2 | Concordant |
| | 14 | 102840084-103639456 | rs12147883 | 5.42E-14 | rs7158166 | 1.13E-06 | 0.07 | 0.78 | TRAF3 | Concordant |
| CD8 STIM | 6 | 30706942-31506854 | rs1265094 | 1.86E-31 | rs2535324 | 8.93E-05 | 0.01 | 0.63 | HCG4 | Concordant |
| | 11 | 61196289-61995513 | rs968567 | 2.42E-41 | rs174544 | 1.83E-16 | 0.31 | 0.69 | FADS1 | Concordant |
| | 11 | 61196289-61995513 | rs968567 | 2.42E-41 | rs7943728 | 1.53E-14 | 0.00 | 1.00 | FADS2 | Concordant |
| M2 | 11 | 61196289-61995513 | rs968567 | 2.42E-41 | rs174538 | 4.33E-14 | 0.10 | 0.90 | FADS2 | Concordant |
| | 16 | 11317993-12117637 | rs113962704 | 1.91E-12 | rs9932684 | 3.77E-05 | 0.11 | 0.52 | AC099489.1 | Concordant |
| | 16 | 11317993-12117637 | rs113962704 | 1.91E-12 | rs9932684 | 8.75E-07 | 0.06 | 0.81 | LITAF | Concordant |
| Monocytes | 7 | 75634687-76433936 | rs55722505 | 8.61E-14 | rs1799237 | 4.20E-11 | 0.31 | 0.69 | ZP3 | Opposite |
| | 7 | 75634687-76433936 | rs55722505 | 8.61E-14 | rs4728708 | 2.59E-12 | 0.22 | 0.78 | SSC4D | Opposite |
| | 11 | 61196289-61995513 | rs968567 | 2.42E-41 | rs7943728 | 1.46E-14 | 0.00 | 1.00 | FADS2 | Concordant |
| | 16 | 11317993-12117637 | rs113962704 | 1.91E-12 | rs72781049 | 1.73E-12 | 0.02 | 0.98 | LITAF | Concordant |
| NK | 11 | 61196289-61995513 | rs968567 | 2.42E-41 | rs7943728 | 3.87E-14 | 0.03 | 0.97 | FADS2 | Concordant |
| TFH | 11 | 61196289-61995513 | rs968567 | 2.42E-41 | rs174574 | 1.69E-06 | 0.13 | 0.67 | TMEM258 | Concordant |
| | 11 | 61196289-61995513 | rs968567 | 2.42E-41 | rs7943728 | 3.96E-19 | 0.00 | 1.00 | FADS2 | Concordant |
| | 14 | 102840084-103639456 | rs12147883 | 5.42E-14 | rs2075771 | 1.71E-05 | 0.07 | 0.71 | TRAF3 | Concordant |
| TH1 | 2 | 102289183-103088777 | rs13427957 | 6.19E-10 | rs13401717 | 1.06E-04 | 0.05 | 0.69 | MIR4772 | Concordant |
| | 11 | 61196289-61995513 | rs968567 | 2.42E-41 | rs174544 | 1.74E-13 | 0.26 | 0.74 | FADS1 | Concordant |
| | 11 | 61196289-61995513 | rs968567 | 2.42E-41 | rs7943728 | 2.250e-17 | 0.00 | 1.00 | FADS2 | Concordant |
| | 12 | 47814851-48614567 | rs7487637 | 9.97E-15 | rs11168256 | 1.57E-04 | 0.08 | 0.59 | HDAC7 | Concordant |
| | 14 | 102840084-103639456 | rs12147883 | 5.42E-14 | rs2075771 | 1.88E-05 | 0.08 | 0.58 | TRAF3 | Concordant |
| TH2 | 1 | 172763639-173563418 | rs7518129 | 1.06E-16 | rs59597894 | 6.26E-05 | 0.09 | 0.50 | TNFSF18 | Opposite |
| | 11 | 61196289-61995513 | rs968567 | 2.416E-41 | rs796587134 | 2.14E-07 | 0.24 | 0.51 | MYRF | Concordant |
| | 11 | 61196289-61995513 | rs968567 | 2.416E-41 | rs7943728 | 2.02E-06 | 0.03 | 0.89 | TMEM258 | Concordant |
| | 11 | 61196289-61995513 | rs968567 | 2.416E-41 | rs174544 | 5.90E-21 | 0.00 | 1.00 | FADS2 | Concordant |
| | 14 | 102840084-103639456 | rs12147883 | 5.42E-14 | rs12878532 | 5.02E-06 | 0.06 | 0.84 | TRAF3 | Concordant |
| TH17 | 11 | 61196289-61995513 | rs968567 | 2.416E-41 | rs174528 | 3.83E-06 | 0.09 | 0.71 | TMEM258 | Concordant |
| | 11 | 61196289-61995513 | rs968567 | 2.416E-41 | rs7943728 | 7.17E-20 | 0.00 | 1.00 | FADS2 | Concordant |
| | 14 | 102840084-103639456 | rs12147883 | 5.416E-14 | rs79589176 | 6.04E-05 | 0.09 | 0.57 | TRAF3 | Concordant |
| | 22 | 30048571-30848388 | rs193473 | 1.58E-20 | rs249398 | 3.58E-05 | 0.15 | 0.52 | CNN2P1 | Opposite |
| THSTAR | 6 | 30706942-31506854 | rs1265094 | 1.858E-31 | rs3130702 | 3.49E-05 | 0.27 | 0.55 | APOM | Concordant |
| | 11 | 61196289-61995513 | rs968567 | 2.416E-41 | rs174576 | 3.63E-17 | 0.36 | 0.64 | FADS1 | Concordant |
| | 11 | 61196289-61995513 | rs968567 | 2.416E-41 | rs7943728 | 1.32E-14 | 0.01 | 0.99 | FADS2 | Concordant |
| | 14 | 102840084-103639456 | rs12147883 | 5.416E-14 | rs2075771 | 4.04E-06 | 0.07 | 0.76 | TRAF3 | Concordant |
| | 22 | 30048571-30848388 | rs193473 | 1.58E-20 | rs199726005 | 3.38E-05 | 0.11 | 0.61 | RF00019 | Concordant |
| TREG MEM | 5 | 55039671-55838571 | rs6859219 | 1.413E-20 | rs7731626 | 3.02E-08 | 0.12 | 0.51 | ANKRD55 | Concordant |
| | 6 | 30706942-31506854 | rs1265094 | 1.858E-31 | rs1265157 | 1.01E-04 | 0.18 | 0.60 | PRRT1 | Opposite |
| | 11 | 61196289-61995513 | rs968567 | 2.416E-41 | rs174576 | 2.69E-18 | 0.22 | 0.78 | FADS1 | Concordant |
| | 11 | 61196289-61995513 | rs968567 | 2.416E-41 | rs7943728 | 6.73E-18 | 0.00 | 1.00 | FADS2 | Concordant |
| | 11 | 61196289-61995513 | rs968567 | 2.416E-41 | rs174585 | 1.64E-10 | 0.15 | 0.83 | FADS3 | Concordant |
| | 14 | 102840084-103639456 | rs12147883 | 5.416E-14 | rs12887521 | 8.63E-06 | 0.08 | 0.76 | TRAF3 | Concordant |
| | 16 | 30516148-31314599 | rs1458201 | 2.017E-11 | rs11865499 | 1.58E-05 | 0.05 | 0.67 | KAT8 | Concordant |
| TREG NAIVE | 11 | 61196289-61995513 | rs968567 | 2.416E-41 | rs7943728 | 4.97E-18 | 0.00 | 1.00 | FADS1 | Concordant |
| | 11 | 61196289-61995513 | rs968567 | 2.416E-41 | rs7943728 | 1.83E-15 | 0.00 | 1.00 | FADS2 | Concordant |
| Whole blood | 5 | 55039671-55838571 | rs6859219 | 1.413E-20 | rs7731626 | 3.27E-310 | 0.02 | 0.98 | ANKRD55 | Concordant |
| | 5 | 94878263-95677384 | rs3777175 | 7.813E-30 | rs1841010 | 1.16E-37 | 0.47 | 0.53 | ELL2 | Opposite |
| | 6 | 30706942-31506854 | rs1265094 | 1.858E-31 | rs114986457 | 9.23E-25 | 0.00 | 1.00 | HCG4 | Concordant |
| | 6 | 30706942-31506854 | rs1265094 | 1.858E-31 | rs145930603 | 2.45E-35 | 0.02 | 0.98 | HCG4P3 | Opposite |
| | 6 | 30706942-31506854 | rs1265094 | 1.858E-31 | rs116481148 | 3.27E-310 | 0.04 | 0.93 | HLA-DRB5 | Concordant |
| | 6 | 30706942-31506854 | rs1265094 | 1.858E-31 | rs116239366 | 1.27E-20 | 0.07 | 0.93 | DDX39BP2 | Opposite |
| | 6 | 30706942-31506854 | rs1265094 | 1.858E-31 | rs114986457 | 1.33E-101 | 0.47 | 0.53 | HLA-K | Concordant |
| | 6 | 45127152-45926460 | rs1200427 | 6.853E-14 | rs1200428 | 3.27E-310 | 0.01 | 0.99 | RUNX2 | Opposite |
| | 7 | 75634687-76433936 | rs55722505 | 8.611E-14 | rs55650029 | 5.74E-42 | 0.04 | 0.96 | SRCRB4D | Opposite |
| | 7 | 75634687-76433936 | rs55722505 | 8.611E-14 | rs6966715 | 3.27E-310 | 0.14 | 0.86 | DTX2 | Opposite |
| | 7 | 75634687-76433936 | rs55722505 | 8.611E-14 | rs4415243 | 3.47E-227 | 0.47 | 0.53 | YWHAG | Opposite |
| | 11 | 61196289-61995513 | rs968567 | 2.416E-41 | rs61896141 | 3.27E-310 | 0.09 | 0.91 | FADS1 | Concordant |
| | 11 | 61196289-61995513 | rs968567 | 2.416E-41 | rs968567 | 3.27E-310 | 0.15 | 0.85 | FADS2 | Concordant |
| | 12 | 111437618-112233495 | rs10774624 | 1.374E-13 | rs597808 | 6.93E-07 | 0.01 | 0.99 | PPTC7 | Opposite |
| | 12 | 111437618-112233495 | rs10774624 | 1.374E-13 | rs138281696 | 2.47E-45 | 0.02 | 0.98 | TRAFD1 | Concordant |
| | 12 | 111437618-112233495 | rs10774624 | 1.374E-13 | rs73197952 | 1.07E-09 | 0.02 | 0.97 | HVCN1 | Opposite |
| | 16 | 30516148-31314599 | rs1458201 | 2.017E-11 | rs4889653 | 3.00E-06 | 0.09 | 0.89 | SRCAP | Concordant |
| | 16 | 30516148-31314599 | rs1458201 | 2.017E-11 | rs17855121 | 1.05E-07 | 0.14 | 0.86 | PPP4C | Opposite |
| | 16 | 30516148-31314599 | rs1458201 | 2.017E-11 | rs4346218 | 9.69E-08 | 0.30 | 0.68 | FBXL19-AS1 | Concordant |

**Supplementary Table 8. Top human orthologs of mouse genes with known mouse knock-out phenotypes of "abnormal IgA levels" (mouse ontology database) and their regional association with IgA levels based on GWAS meta-analysis.** The columns include chromosome number, start and end locations of the gene region, strand information for each gene, gene name, and the smallest P value within each gene region. The results meeting Bonferroni-corrected threshold (*P* < 0.05 / 22169 = 2.26E-6) are listed, 11 of 13 prioritized genes reside within the genome-wide significant loci.

| CHR | START | STOP | SIZE | STRAND | GENE | P |
|---|---|---|---|---|---|---|
| 1 | 25226001 | 25291501 | 65500 | - | *RUNX3* | 3.42E-92 |
| 5 | 95220801 | 95297775 | 76974 | - | *ELL2* | 7.81E-30 |
| 6 | 31539875 | 31542100 | 2225 | + | *LTA* | 7.15E-26 |
| 6 | 31548335 | 31550202 | 1867 | - | *LTB* | 8.60E-24 |
| 6 | 31543343 | 31546112 | 2769 | + | *TNF* | 8.60E-24 |
| 11 | 65421066 | 65430443 | 9377 | - | *RELA* | 2.57E-22 |
| 5 | 55230924 | 55290821 | 59897 | - | *IL6ST* | 1.41E-20 |
| 11 | 111222980 | 111250157 | 27177 | - | *POU2AF1* | 7.01E-16 |
| 14 | 103243815 | 103377837 | 134022 | + | *TRAF3* | 5.42E-14 |
| 16 | 31212806 | 31214097 | 1291 | - | *PYCARD* | 2.02E-11 |
| 6 | 33540322 | 33548070 | 7748 | - | *BAK1* | 2.92E-11 |
| 17 | 7461608 | 7464925 | 3317 | + | *TNFSF13* | 9.41E-08 |
| 2 | 61108629 | 61155291 | 46662 | + | *REL* | 1.52E-06 |

**Supplementary Table 9. Top human orthologs of mouse genes with known mouse knock-out phenotypes of "abnormal immune tolerance" (mouse ontology database) and their regional association with IgA levels based on GWAS meta-analysis.** The columns include chromosome number, start and end locations of the gene region, strand information for each gene, gene name, and the smallest P value within each gene region. The results meeting Bonferroni-corrected threshold (*P* < 0.05 / 22169 = 2.26E-6) are listed.

| CHR | START | STOP | SIZE | STRAND | GENE | P |
|---|---|---|---|---|---|---|
| 11 | 61560108 | 61564714 | 4606 | + | *FEN1* | 2.42E-41 |
| 11 | 61281187 | 61348344 | 67157 | - | *SYT7* | 2.42E-41 |
| 6 | 32158542 | 32163300 | 4758 | - | *GPSM3* | 9.70E-27 |
| 6 | 32627240 | 32634466 | 7226 | - | *HLA-DQB1* | 9.70E-27 |
| 6 | 32812985 | 32821748 | 8763 | - | *TAP1* | 1.90E-26 |
| 6 | 32916390 | 32920899 | 4509 | - | *HLA-DMA* | 6.78E-26 |
| 6 | 31539875 | 31542100 | 2225 | + | *LTA* | 7.15E-26 |
| 6 | 29910246 | 29913661 | 3415 | + | *HLA-A* | 9.93E-26 |
| 6 | 29624757 | 29640149 | 15392 | + | *MOG* | 9.93E-26 |
| 6 | 30457182 | 30461982 | 4800 | + | *HLA-E* | 1.54E-24 |
| 6 | 31582993 | 31584798 | 1805 | + | *AIF1* | 8.60E-24 |
| 6 | 31543343 | 31546112 | 2769 | + | *TNF* | 8.60E-24 |
| 6 | 31949833 | 31970457 | 20624 | + | *C4A* | 4.97E-23 |
| 6 | 31949833 | 31970458 | 20625 | + | *C4B* | 4.97E-23 |
| 6 | 31913720 | 31919861 | 6141 | + | *CFB* | 7.97E-22 |
| 5 | 55230924 | 55290821 | 59897 | - | *IL6ST* | 1.41E-20 |
| 9 | 117546914 | 117568408 | 21494 | - | *TNFSF15* | 1.13E-18 |
| 1 | 173152869 | 173176471 | 23602 | - | *TNFSF4* | 1.06E-16 |
| 11 | 111222980 | 111250157 | 27177 | - | *POU2AF1* | 7.01E-16 |
| 12 | 48103517 | 48119355 | 15838 | - | *ENDOU* | 9.97E-15 |
| 12 | 48436680 | 48500091 | 63411 | - | *SENP1* | 9.97E-15 |
| 12 | 48235319 | 48298814 | 63495 | - | *VDR* | 9.97E-15 |
| 14 | 103243815 | 103377837 | 134022 | + | *TRAF3* | 5.42E-14 |
| 16 | 11348273 | 11350039 | 1766 | - | *SOCS1* | 1.91E-12 |
| 6 | 33540322 | 33548070 | 7748 | - | *BAK1* | 2.92E-11 |
| 2 | 102686835 | 102796334 | 109499 | + | *IL1R1* | 6.19E-10 |
| 17 | 7387697 | 7417935 | 30238 | + | *POLR2A* | 9.41E-08 |
| 17 | 7461608 | 7464925 | 3317 | + | *TNFSF13* | 9.41E-08 |
| 2 | 61108629 | 61155291 | 46662 | + | *REL* | 1.52E-06 |

**Supplementary Table 10. Top human orthologs of mouse genes with known mouse knock-out phenotype of "abnormal response to infection" (mouse ontology database) and their regional association with IgA levels based on GWAS meta-analysis.** The columns include chromosome number, start and end locations of the gene region, strand information for each gene, gene name, and the smallest P value within each gene region. The results meeting Bonferroni-corrected threshold (*P* < 0.05 / 22169 = 2.26E-6) are listed.

| CHR | START | STOP | SIZE | STRAND | GENE | P |
|---|---|---|---|---|---|---|
| 1 | 25071759 | 25170815 | 99056 | + | *CLIC4* | 3.42E-92 |
| 6 | 30710975 | 30712327 | 1352 | - | *IER3* | 1.86E-31 |
| 6 | 32780539 | 32784825 | 4286 | - | *HLA-DOB* | 9.70E-27 |
| 6 | 32627240 | 32634466 | 7226 | - | *HLA-DQB1* | 9.70E-27 |
| 6 | 32821937 | 32827628 | 5691 | + | *PSMB9* | 1.90E-26 |
| 6 | 32812985 | 32821748 | 8763 | - | *TAP1* | 1.90E-26 |
| 6 | 32916390 | 32920899 | 4509 | - | *HLA-DMA* | 6.78E-26 |
| 6 | 31539875 | 31542100 | 2225 | + | *LTA* | 7.15E-26 |
| 6 | 29910246 | 29913661 | 3415 | + | *HLA-A* | 9.93E-26 |
| 6 | 29794755 | 29798899 | 4144 | + | *HLA-G* | 9.93E-26 |
| 6 | 29523388 | 29527702 | 4314 | - | *UBD* | 9.93E-26 |
| 6 | 30457182 | 30461982 | 4800 | + | *HLA-E* | 1.54E-24 |
| 6 | 31548335 | 31550202 | 1867 | - | *LTB* | 8.60E-24 |
| 6 | 31543343 | 31546112 | 2769 | + | *TNF* | 8.60E-24 |
| 6 | 31949833 | 31970457 | 20624 | + | *C4A* | 4.97E-23 |
| 6 | 31949833 | 31970458 | 20625 | + | *C4B* | 4.97E-23 |
| 11 | 65365225 | 65381720 | 16495 | - | *MAP3K11* | 2.57E-22 |
| 11 | 65421066 | 65430443 | 9377 | - | *RELA* | 2.57E-22 |
| 6 | 31913720 | 31919861 | 6141 | + | *CFB* | 7.97E-22 |
| 6 | 31847535 | 31865484 | 17949 | - | *EHMT2* | 2.08E-21 |
| 5 | 55147206 | 55218682 | 71476 | + | *IL31RA* | 1.41E-20 |
| 5 | 55230924 | 55290821 | 59897 | - | *IL6ST* | 1.41E-20 |
| 22 | 30636435 | 30642840 | 6405 | - | *LIF* | 1.58E-20 |
| 11 | 111222980 | 111250157 | 27177 | - | *POU2AF1* | 7.01E-16 |
| 1 | 27189632 | 27190947 | 1315 | + | *SFN* | 9.94E-15 |
| 12 | 48235319 | 48298814 | 63495 | - | *VDR* | 9.97E-15 |
| 16 | 11348273 | 11350039 | 1766 | - | *SOCS1* | 1.91E-12 |
| 16 | 31271287 | 31344213 | 72926 | + | *ITGAM* | 2.02E-11 |
| 16 | 31212806 | 31214097 | 1291 | - | *PYCARD* | 2.02E-11 |
| 2 | 102686835 | 102796334 | 109499 | + | *IL1R1* | 6.19E-10 |
| 2 | 102927961 | 102968497 | 40536 | + | *IL1RL1* | 6.19E-10 |
| 9 | 117096432 | 117156685 | 60253 | - | *AKNA* | 2.22E-09 |
| 17 | 7076750 | 7082883 | 6133 | - | *ASGR1* | 9.41E-08 |
| 17 | 7743234 | 7758118 | 14884 | + | *KDM6B* | 9.41E-08 |
| 17 | 7452374 | 7461207 | 8833 | + | *TNFSF12* | 9.41E-08 |
| 13 | 50571142 | 50592603 | 21461 | + | *TRIM13* | 2.86E-07 |
| 19 | 55417507 | 55424439 | 6932 | + | *NCR1* | 5.01E-07 |
| 7 | 128577990 | 128590096 | 12106 | + | *IRF5* | 1.06E-06 |
| 1 | 27938800 | 27961727 | 22927 | - | *FGR* | 1.23E-06 |
| 2 | 143886898 | 144525921 | 639023 | + | *ARHGAP15* | 1.50E-06 |
| 2 | 61108629 | 61155291 | 46662 | + | *REL* | 1.52E-06 |

**Supplementary Table 11. The associations of top SNPs from GWAS for IgA levels in GWAS for IgA nephropathy.** Among 31 independent alleles associated with increased IgA levels, 12 had nominal association with increased risk of IgA nephropathy at *P*<0.05 (all with concordant effects).

| Locus | CHR | BP | SNP | IgA level GWAS | | | IgA Nephropathy GWAS | | | Direction |
|---|---|---|---|---|---|---|---|---|---|---|
| | | | | Risk Allele | BETA | P-value | Risk Allele | BETA | P-value | |
| *RUNX3* | 1 | 25291697 | rs188468174 | C | 0.876 | 3.42E-92 | C | 0.009 | 9.68E-01 | - |
| *TNFSF4* | 1 | 173163568 | rs7518129 | G | 0.056 | 1.06E-16 | G | 0.118 | 6.71E-07 | Concordant |
| | 1 | 173172158 | rs4916314 | A | 0.061 | 3.46E-14 | A | 0.147 | 9.09E-07 | Concordant |
| *IL1R1* | 2 | 102689031 | rs13427957 | C | 0.040 | 6.19E-10 | C | 0.024 | 2.48E-01 | - |
| *ELL2* | 5 | 95277555 | rs3777175 | G | 0.084 | 7.81E-30 | A | 0.005 | 8.73E-01 | - |
| | 5 | 95231115 | rs3777207 | T | 0.080 | 1.16E-25 | C | 0.039 | 3.23E-01 | - |
| | 5 | 95286609 | rs200990904 | C | 0.065 | 2.02E-14 | T | 0.007 | 8.31E-01 | - |
| *ANKRD55* | 5 | 55438580 | rs6859219 | C | 0.073 | 1.41E-20 | C | 0.167 | 1.96E-07 | Concordant |
| *HLA* | 6 | 31106893 | rs1265094 | A | 0.076 | 1.86E-31 | A | 0.047 | 2.70E-02 | Concordant |
| | 6 | 32710119 | rs5018343 | A | 0.070 | 2.94E-24 | T | 0.001 | 9.63E-01 | - |
| | 6 | 32449523 | chr6:32449523 | T | 0.078 | 4.32E-25 | C | 0.012 | 0.826 | - |
| | 6 | 29943715 | rs3823363 | C | 0.062 | 3.13E-22 | C | 0.068 | 2.88E-04 | Concordant |
| | 6 | 31438243 | rs2596450 | G | 0.058 | 1.33E-17 | A | 0.014 | 6.43E-01 | - |
| | 6 | 32804299 | rs3819717 | C | 0.047 | 1.01E-13 | T | 0.028 | 1.62E-01 | - |
| | 6 | 31324152 | rs1050628 | G | 0.126 | 3.56E-16 | A | 0.085 | 5.29E-01 | - |
| | 6 | 32343369 | rs3117106 | T | 0.073 | 1.44E-12 | T | 0.305 | 7.39E-20 | Concordant |
| *RUNX2* | 6 | 45526470 | rs1200427 | T | 0.059 | 6.85E-14 | T | 0.045 | 2.32E-01 | - |
| *FILNC1* | 6 | 139975943 | rs17069163 | T | 0.050 | 5.94E-11 | C | 0.007 | 7.68E-01 | - |
| *ZP3,SSC4D* | 7 | 76034150 | rs55722505 | C | 0.048 | 8.61E-14 | C | 0.034 | 1.80E-01 | - |
| *TNFSF8* | 9 | 117692882 | rs3181356 | T | 0.069 | 1.13E-18 | T | 0.163 | 1.03E-04 | Concordant |
| *FADS2* | 11 | 61595564 | rs968567 | T | 0.116 | 2.42E-41 | T | 0.050 | 1.21E-01 | - |
| | 11 | 61557803 | rs102275 | C | 0.073 | 3.60E-29 | C | 0.006 | 7.79E-01 | - |
| *OVOL1* | 11 | 65555524 | rs10896045 | A | 0.066 | 2.57E-22 | A | 0.168 | 4.77E-13 | Concordant |
| *POU2AF1* | 11 | 111267394 | rs4938518 | T | 0.056 | 7.01E-16 | T | 0.016 | 4.66E-01 | - |
| *HDAC7* | 12 | 48214825 | rs7487637 | G | 0.054 | 9.97E-15 | G | 0.100 | 6.78E-03 | Concordant |
| *SH2B3* | 12 | 111833788 | rs10774624 | G | 0.050 | 1.37E-13 | G | 0.092 | 1.23E-02 | Concordant |
| *RCOR1,TRAF3* | 14 | 103239630 | rs12147883 | C | 0.048 | 5.42E-14 | C | 0.042 | 6.40E-02 | - |
| *LITAF* | 16 | 11717832 | rs113962704 | T | 0.054 | 1.91E-12 | T | 0.055 | 1.96E-01 | - |
| *CTF1* | 16 | 30916129 | rs1458201 | A | 0.052 | 2.02E-11 | A | 0.019 | 4.61E-01 | - |
| *HORMAD2* | 22 | 30448399 | rs193473 | A | 0.067 | 1.58E-20 | A | 0.139 | 9.07E-07 | Concordant |
| | 22 | 30269907 | rs9614090 | A | 0.052 | 5.56E-16 | A | 0.144 | 1.49E-07 | Concordant |

**Supplementary Table 12. Colocalization analysis of GWAS loci for IgA levels with GWAS signals for IgA nephropathy** PP1: probability of the locus being associated with only IgA levels; PP2: probability of the locus being associated with only IgA nephropathy; PP3: probability of not sharing the same casual variant at the locus; and PP4: probability of sharing the same causal variant at the locus between the two traits. High probability of sharing causal variants was evident for five loci (*).

| Locus | #SNPs | CHR | Start | End | PP1 | PP2 | PP3 | PP4 |
|---|---|---|---|---|---|---|---|---|
| *OVOL1,RELA* | 2013 | 11 | 65156514 | 65955511 | 0.00 | 0.00 | 0.00 | 1.00* |
| *ANKRD55,IL6ST* | 2804 | 5 | 55039671 | 55838485 | 0.00 | 0.00 | 0.00 | 1.00* |
| *HORMAD2,LIF* | 2224 | 22 | 30048571 | 30848388 | 0.00 | 0.00 | 0.02 | 0.98* |
| *TNFSF4,TNFSF18* | 2021 | 1 | 172763639 | 173563418 | 0.00 | 0.00 | 0.04 | 0.96* |
| *SH2B3* | 1031 | 12 | 111438001 | 112233495 | 0.09 | 0.00 | 0.02 | 0.90* |
| *HLA* | 15091 | 6 | 30706942 | 31506854 | 0.00 | 0.00 | 1.00 | 0.00 |
| *CTF1* | 819 | 16 | 30516148 | 31314599 | 0.01 | 0.00 | 0.99 | 0.00 |
| *RCOR1,TRAF3* | 2415 | 14 | 102840160 | 103639456 | 0.03 | 0.00 | 0.89 | 0.08 |
| *TNFSF8* | 3254 | 9 | 117293051 | 118092515 | 0.01 | 0.00 | 0.83 | 0.16 |
| *RUNX2* | 2826 | 6 | 45127152 | 45926460 | 0.38 | 0.00 | 0.61 | 0.01 |
| *LITAF* | 3543 | 16 | 11317993 | 12117447 | 0.85 | 0.00 | 0.14 | 0.02 |
| *POU2AF1* | 2328 | 11 | 110867986 | 111666128 | 0.86 | 0.00 | 0.13 | 0.01 |
| *IL1R1* | 2715 | 2 | 102289430 | 103088777 | 0.87 | 0.00 | 0.12 | 0.02 |
| *RUNX3* | 1953 | 1 | 24891715 | 25689406 | 0.85 | 0.00 | 0.11 | 0.03 |
| *FILNC1* | 2459 | 6 | 139576015 | 140375356 | 0.90 | 0.00 | 0.09 | 0.01 |
| *FADS2,FADS1* | 2288 | 11 | 61196289 | 61995513 | 0.89 | 0.00 | 0.08 | 0.03 |
| *HDAC7, VDR* | 3153 | 12 | 47814851 | 48614567 | 0.72 | 0.00 | 0.06 | 0.22 |
| *ELL2* | 1753 | 5 | 94878263 | 95677384 | 0.94 | 0.00 | 0.05 | 0.01 |
| *ZP3,SSC4D* | 2200 | 7 | 75634687 | 76433936 | 0.94 | 0.00 | 0.05 | 0.02 |

**Supplementary Table 13. Colocalization analysis of GWAS loci for IgA levels with GWAS signals for tonsillectomy**
PP1: probability of the locus being associated with only IgA levels; PP2: probability of the locus being associated with only tonsillectomy; PP3: probability of not sharing the same casual variant at the locus; and PP4: probability of sharing the same causal variant at the locus between the two traits. High probability of sharing causal variants was evident for two loci (*).

| Locus | #SNPs | CHR | Start | End | PP1 | PP2 | PP3 | PP4 |
|---|---|---|---|---|---|---|---|---|
| *SH2B3* | 1542 | 12 | 111438001 | 112233495 | 0.00 | 0.00 | 0.01 | 0.99* |
| *HORMAD2,LIF* | 2494 | 22 | 30048571 | 30848388 | 0.00 | 0.00 | 0.22 | 0.78* |
| *HLA* | 13524 | 6 | 30706942 | 31506854 | 0.00 | 0.00 | 1.00 | 0.00 |
| *CTF1* | 1119 | 16 | 30516148 | 31314599 | 0.09 | 0.00 | 0.91 | 0.00 |
| *RCOR1,TRAF3* | 2307 | 14 | 102840084 | 103639456 | 0.52 | 0.00 | 0.47 | 0.01 |
| *LITAF* | 3549 | 16 | 11317993 | 12117566 | 0.58 | 0.00 | 0.42 | 0.00 |
| *IL1R1* | 2920 | 2 | 102289430 | 103088777 | 0.50 | 0.00 | 0.39 | 0.11 |
| *FILNC1* | 2607 | 6 | 139576015 | 140374954 | 0.83 | 0.00 | 0.16 | 0.00 |
| *TNFSF8* | 3268 | 9 | 117293051 | 118092515 | 0.90 | 0.00 | 0.08 | 0.02 |
| *TNFSF4,TNFSF18* | 2430 | 1 | 172763639 | 173563418 | 0.93 | 0.00 | 0.06 | 0.01 |
| *FADS2,FADS1* | 2581 | 11 | 61196289 | 61995513 | 0.94 | 0.00 | 0.05 | 0.00 |
| *POU2AF1* | 2516 | 11 | 110867986 | 111666128 | 0.93 | 0.00 | 0.05 | 0.02 |
| *RUNX3* | 2213 | 1 | 24891715 | 25688276 | 0.93 | 0.00 | 0.04 | 0.02 |
| *HDAC7, VDR* | 3198 | 12 | 47814851 | 48614567 | 0.96 | 0.00 | 0.04 | 0.00 |
| *OVOL1,RELA* | 2273 | 11 | 65156283 | 65955511 | 0.96 | 0.00 | 0.04 | 0.00 |
| *ANKRD55,IL6ST* | 2964 | 5 | 55039671 | 55838571 | 0.96 | 0.00 | 0.03 | 0.00 |
| *RUNX2* | 2971 | 6 | 45127152 | 45926460 | 0.97 | 0.00 | 0.03 | 0.00 |
| *ELL2* | 2170 | 5 | 94878263 | 95677384 | 0.97 | 0.00 | 0.02 | 0.00 |
| *ZP3,SSC4D* | 2082 | 7 | 75634687 | 76433287 | 0.98 | 0.00 | 0.02 | 0.00 |

**Supplementary Table 14. Genome-wide genetic correlations between serum IgA level and related traits or disorders without HLA region (left) and with HLA region (right).** All analyses are performed with LDSC software, h2: SNP-based heritability, rg: genetic correlation coefficient, se: standard error, z: z-score, p: p-value.

| Trait | Reference | h2 | Correlations with HLA region | | | | Correlations without HLA region | | | |
|---|---|---|---|---|---|---|---|---|---|---|
| | | | rg | se | z | p | rg | se | z | p |
| IgA deficiency | Bronson et al. Nat Genet 2016 | 1.000 | -0.38 | 0.10 | 3.67 | 2.00E-04 | 0.20 | 0.10 | 2.12 | 3.44E-02 |
| Celiac Disease | Zhou et al. Nat Genet 2018 | 0.054 | -0.27 | 0.05 | -5.60 | 2.16E-08 | -0.21 | 0.08 | -2.71 | 6.80E-03 |
| Primary Sclerosing Cholangitis | Ji et al. Nat Genet 2017 | 0.682 | -0.27 | 0.15 | -1.77 | 7.74E-02 | -0.16 | 0.08 | -2.01 | 4.46E-02 |
| Membranous Nephropathy | Xie et al. Nat Comm 2020 | 0.600 | -0.20 | 0.07 | -2.96 | 3.10E-03 | -0.14 | 0.15 | -0.99 | 3.24E-01 |
| Crohn’s Disease | De Lange et al. Nat Genet 2017 | 0.261 | -0.15 | 0.06 | -2.55 | 1.09E-02 | -0.19 | 0.07 | -2.79 | 5.30E-03 |
| Type 1 Diabetes | Zhou et al. Nat Genet 2018 | 0.011 | -0.13 | 0.23 | -0.57 | 5.69E-01 | -0.07 | 0.16 | -0.46 | 6.42E-01 |
| Systemic Lupus Erythematosus | Bentham et al. Nat Genet 2015 | 0.491 | -0.12 | 0.13 | -0.88 | 3.76E-01 | -0.04 | 0.09 | -0.40 | 6.89E-01 |
| Inflammatory Bowel Diseases | De Lange et al. Nat Genet 2017 | 0.183 | -0.12 | 0.05 | -2.23 | 2.58E-02 | -0.13 | 0.06 | -2.08 | 3.76E-02 |
| Ulcerative Colitis | De Lange et al. Nat Genet 2017 | 0.165 | -0.06 | 0.07 | -0.90 | 3.70E-01 | -0.03 | 0.08 | -0.41 | 6.80E-01 |
| Allergy | Ferreira et al. Nat Genet 2017 | 0.031 | 0.08 | 0.04 | 1.95 | 5.06E-02 | 0.07 | 0.06 | 1.14 | 2.55E-01 |
| Rheumatoid Arthritis | Okada et al. Nature 2014 | 0.344 | 0.11 | 0.08 | 1.33 | 1.83E-01 | 0.15 | 0.06 | 2.41 | 1.58E-02 |
| Multiple Sclerosis | Int. MS Genetics Consortium. Science 2019 | 0.250 | 0.24 | 0.09 | 2.75 | 5.90E-03 | 0.05 | 0.17 | 0.28 | 7.83E-01 |
| IgA Nephropathy | Kiryluk et al. accomplishing paper | 0.047 | 0.31 | 0.10 | 3.08 | 2.10E-03 | 0.35 | 0.11 | 3.07 | 2.10E-03 |
| bacterial meningitis | Tian et al. Nat Comm 2017 | 0.003 | -0.35 | 0.13 | -2.65 | 8.10E-03 | -0.47 | 0.16 | -2.82 | 4.80E-03 |
| Positive TB Test | Tian et al. Nat Comm 2017 | 0.009 | -0.32 | 0.28 | -1.15 | 2.52E-01 | -0.17 | 0.30 | -0.58 | 5.61E-01 |
| Shingles | Tian et al. Nat Comm 2017 | 0.013 | -0.23 | 0.11 | -1.99 | 4.71E-02 | -0.46 | 0.17 | -2.63 | 8.60E-03 |
| Measles | Tian et al. Nat Comm 2017 | 0.002 | -0.22 | 0.65 | -0.34 | 7.34E-01 | -0.20 | 0.22 | -0.92 | 3.57E-01 |
| Rubella | Tian et al. Nat Comm 2017 | 0.005 | -0.22 | 0.10 | -2.28 | 2.29E-02 | -0.27 | 0.17 | -1.59 | 1.12E-01 |
| Scarlet fever | Tian et al. Nat Comm 2017 | 0.007 | -0.19 | 0.13 | -1.47 | 1.43E-01 | -0.11 | 0.13 | -0.84 | 4.00E-01 |
| Mononucleosis | Tian et al. Nat Comm 2017 | 0.004 | -0.16 | 0.29 | -0.57 | 5.71E-01 | -0.54 | 0.31 | -1.73 | 8.41E-02 |
| Mumps | Tian et al. Nat Comm 2017 | 0.005 | -0.11 | 0.16 | -0.72 | 4.72E-01 | 0.01 | 0.17 | 0.07 | 9.42E-01 |
| Cold sores | Tian et al. Nat Comm 2017 | 0.005 | -0.08 | 0.26 | -0.31 | 7.57E-01 | 0.15 | 0.22 | 0.70 | 4.84E-01 |
| Myringotomy | Tian et al. Nat Comm 2017 | 0.006 | -0.07 | 0.09 | -0.71 | 4.75E-01 | -0.01 | 0.13 | -0.08 | 9.38E-01 |
| Chicken pox | Tian et al. Nat Comm 2017 | 0.008 | -0.06 | 0.08 | -0.76 | 4.46E-01 | 0.01 | 0.12 | 0.09 | 9.25E-01 |
| Hepatitis B | Tian et al. Nat Comm 2017 | 0.006 | -0.02 | 0.13 | -0.14 | 8.86E-01 | 0.20 | 0.12 | 1.70 | 8.95E-02 |
| Childhood Ear Infections | Tian et al. Nat Comm 2017 | 0.018 | -0.01 | 0.07 | -0.15 | 8.79E-01 | 0.06 | 0.09 | 0.63 | 5.26E-01 |
| Yeast infections | Tian et al. Nat Comm 2017 | 0.009 | 0.00 | 0.07 | 0.07 | 9.45E-01 | 0.01 | 0.09 | 0.10 | 9.20E-01 |
| UTI Frequency | Tian et al. Nat Comm 2017 | 0.011 | 0.01 | 0.08 | 0.10 | 9.19E-01 | -0.02 | 0.10 | -0.15 | 8.78E-01 |
| Colds Last Year | Tian et al. Nat Comm 2017 | 0.008 | 0.02 | 0.08 | 0.23 | 8.17E-01 | -0.03 | 0.11 | -0.26 | 7.95E-01 |
| Rheumatic Fever | Tian et al. Nat Comm 2017 | 0.004 | 0.03 | 0.14 | 0.20 | 8.39E-01 | 0.19 | 0.15 | 1.27 | 2.04E-01 |
| Strep Throat | Tian et al. Nat Comm 2017 | 0.007 | 0.04 | 0.20 | 0.18 | 8.60E-01 | 0.24 | 0.15 | 1.61 | 1.07E-01 |
| Tonsillectomy | Tian et al. Nat Comm 2017 | 0.028 | 0.08 | 0.11 | 0.75 | 4.56E-01 | 0.20 | 0.08 | 2.48 | 1.31E-02 |
| Plantar Warts | Tian et al. Nat Comm 2017 | 0.014 | 0.09 | 0.27 | 0.33 | 7.39E-01 | -0.24 | 0.12 | -1.96 | 4.95E-02 |
| Hepatitis A | Tian et al. Nat Comm 2017 | 0.003 | 0.12 | 0.21 | 0.54 | 5.86E-01 | -0.06 | 0.17 | -0.36 | 7.19E-01 |
| Chronic Sinus Infections | Tian et al. Nat Comm 2017 | 0.004 | 0.12 | 0.11 | 1.12 | 2.62E-01 | 0.05 | 0.15 | 0.35 | 7.28E-01 |
| Pneumonia | Tian et al. Nat Comm 2017 | 0.014 | 0.18 | 0.11 | 1.63 | 1.03E-01 | 0.08 | 0.11 | 0.77 | 4.44E-01 |
| Serum LDL | Willer et al. Nat Gen 2013 | 0.035 | -0.10 | 0.07 | -1.44 | 1.49E-01 | -0.14 | 0.09 | -1.51 | 1.32E-01 |
| Serum Cholesterol | Willer et al. Nat Gen 2013 | 0.047 | -0.09 | 0.05 | -1.73 | 8.41E-02 | -0.17 | 0.09 | -1.83 | 6.78E-02 |
| Height | Wood et al. Nat Genet 2014 | 0.413 | -0.05 | 0.04 | -1.09 | 2.77E-01 | -0.03 | 0.05 | -0.54 | 5.87E-01 |
| Serum Triglycerides | Willer et al. Nat Gen 2013 | 0.054 | -0.02 | 0.10 | -0.16 | 8.71E-01 | 0.03 | 0.11 | 0.22 | 8.27E-01 |
| eGFR Creatinine | Wuttke et al. Nat Genet 2019 | 0.062 | 0.02 | 0.07 | 0.35 | 7.29E-01 | 0.01 | 0.09 | 0.14 | 8.89E-01 |
| eGFR Cystatine C | Wuttke et al. Nat Genet 2019 | 0.086 | 0.02 | 0.09 | 0.27 | 7.90E-01 | 0.06 | 0.13 | 0.48 | 6.34E-01 |
| Coronary Artery Disease | Nikpay et al. Nat Genet 2015 | 0.040 | 0.05 | 0.04 | 1.27 | 2.02E-01 | 0.08 | 0.05 | 1.73 | 8.36E-02 |
| Essential Hypertension | Zhou et al. Nat Genet 2018 | 0.033 | 0.05 | 0.05 | 1.12 | 2.65E-01 | 0.09 | 0.05 | 1.77 | 7.74E-02 |
| Albuminuria | Haas et al. Am J Hum Genet 2018 | 0.024 | 0.07 | 0.05 | 1.63 | 1.03E-01 | 0.03 | 0.06 | 0.54 | 5.90E-01 |
| Chronic Kidney Disease | Wuttke et al. Nat Genet 2019 | 0.012 | 0.11 | 0.16 | 0.69 | 4.90E-01 | 0.20 | 0.32 | 0.62 | 5.35E-01 |
| Body Mass Index | Locke et al. Nature 2015 | 0.163 | 0.13 | 0.06 | 2.38 | 1.71E-02 | 0.13 | 0.06 | 2.15 | 3.16E-02 |
| Type 2 Diabetes | Xue et al. Nat Comm 1018 | 0.026 | 0.17 | 0.07 | 2.55 | 1.07E-02 | 0.18 | 0.07 | 2.52 | 1.16E-02 |

**Supplementary Table 15. Meta-phenome-wide association study (Meta-PheWAS) for the genome-wide polygenic score (GPS) for serum IgA levels across eMERGE-III and UKBB datasets.** Only the top associations exceeding the phenome-wide significance threshold are listed for the GPS **a)** with and **b)** without the HLA region. OR: odds ratio per standard deviation of the GPS; SE: standard error; all associations are adjusted for age, sex, site/cohort, genotyping batch, and ancestry, eEMRGE-III participants included in the GWAS for IgA levels were excluded from the analysis.

**a) GPS with HLA**

| Phenotype | Description | Phenotype Group | Beta | OR | SE | P value | N cases | N controls |
|---|---|---|---|---|---|---|---|---|
| 557.1 | Celiac disease | digestive | -0.621 | 0.537 | 0.019 | 4.62E-227 | 2623 | 385965 |
| 244.4 | Hypothyroidism NOS | endocrine/metabolic | -0.060 | 0.941 | 0.007 | 1.77E-19 | 29802 | 460255 |
| 275.1 | Disorders of iron metabolism | hematopoietic | 0.273 | 1.313 | 0.030 | 3.97E-19 | 1333 | 493272 |
| 244 | Hypothyroidism | endocrine/metabolic | -0.058 | 0.944 | 0.007 | 1.03E-18 | 31543 | 460255 |
| 242 | Thyrotoxicosis with or without goiter | endocrine/metabolic | -0.125 | 0.882 | 0.017 | 1.55E-13 | 4473 | 460255 |
| 250.1 | Type 1 diabetes | endocrine/metabolic | -0.093 | 0.911 | 0.014 | 8.22E-12 | 7497 | 447124 |
| 696 | Psoriasis and related disorders | dermatologic | -0.097 | 0.908 | 0.015 | 1.69E-10 | 5369 | 465843 |
| 242.1 | Graves' disease | endocrine/metabolic | -0.202 | 0.817 | 0.032 | 1.74E-10 | 1316 | 460255 |
| 696.4 | Psoriasis | dermatologic | -0.098 | 0.907 | 0.015 | 2.32E-10 | 4996 | 465843 |
| 593 | Hematuria | genitourinary | 0.040 | 1.041 | 0.007 | 1.04E-08 | 26620 | 440047 |
| 250.13 | Type 1 diabetes with ophthalmic manifestations | endocrine/metabolic | -0.184 | 0.832 | 0.033 | 3.18E-08 | 1106 | 447124 |
| 695.3 | Rosacea | dermatologic | 0.132 | 1.141 | 0.024 | 4.45E-08 | 2666 | 474313 |
| 555 | Inflammatory bowel disease and other gastroenteritis and colitis | digestive | 0.068 | 1.071 | 0.013 | 5.83E-08 | 7671 | 385965 |
| 696.41 | Psoriasis vulgaris | dermatologic | -0.093 | 0.912 | 0.017 | 1.01E-07 | 4014 | 465843 |
| 335 | Multiple sclerosis | neurological | 0.119 | 1.126 | 0.024 | 4.37E-07 | 2134 | 460061 |
| 454.1 | Varicose veins of lower extremity | circulatory system | 0.041 | 1.042 | 0.008 | 6.63E-07 | 17180 | 422020 |
| 555.2 | Ulcerative colitis | digestive | 0.073 | 1.076 | 0.015 | 1.41E-06 | 5187 | 385965 |
| 454 | Varicose veins | circulatory system | 0.039 | 1.040 | 0.008 | 1.54E-06 | 18258 | 422020 |
| 250.11 | Type 1 diabetes with ketoacidosis | endocrine/metabolic | -0.230 | 0.794 | 0.048 | 1.75E-06 | 523 | 447124 |
| 695.4 | Lupus (localized and systemic) | dermatologic | -0.143 | 0.867 | 0.030 | 2.51E-06 | 1592 | 471987 |
| 555.21 | Ulcerative colitis (chronic) | digestive | 0.141 | 1.151 | 0.031 | 4.78E-06 | 1415 | 385965 |
| 251.1 | Hypoglycemia | endocrine/metabolic | -0.101 | 0.904 | 0.023 | 7.12E-06 | 2511 | 434768 |
| 695.42 | Systemic lupus erythematosus | dermatologic | -0.146 | 0.864 | 0.033 | 7.27E-06 | 1402 | 471987 |
| 250 | Diabetes mellitus | endocrine/metabolic | -0.025 | 0.975 | 0.006 | 7.68E-06 | 47177 | 447124 |
| 706 | Diseases of sebaceous glands | dermatologic | 0.037 | 1.037 | 0.008 | 8.68E-06 | 21288 | 474164 |
| 580.12 | Non-proliferative glomerulonephritis | genitourinary | -0.238 | 0.788 | 0.054 | 9.63E-06 | 556 | 457242 |
| 695.21 | Dermatitis herpetiformis | dermatologic | -0.550 | 0.577 | 0.124 | 9.97E-06 | 69 | 474313 |
| 285 | Other anemias | hematopoietic | -0.028 | 0.972 | 0.007 | 1.47E-05 | 37100 | 444319 |
| 371 | Inflammation of the eye | sense organs | 0.055 | 1.057 | 0.013 | 1.92E-05 | 9889 | 473293 |
| 250.12 | Type 1 diabetes with renal manifestations | endocrine/metabolic | -0.212 | 0.809 | 0.050 | 2.17E-05 | 599 | 447124 |
| 740 | Osteoarthrosis | musculoskeletal | -0.018 | 0.982 | 0.004 | 3.00E-05 | 81698 | 418302 |

**b) GPS without HLA**

| Phenotype | Description | Phenotype Group | Beta | OR | SE | P value | N cases | N controls |
|---|---|---|---|---|---|---|---|---|
| 557.1 | Celiac disease | digestive | -0.153 | 0.858 | 0.022 | 5.90E-12 | 388588 | 2623 |
| 278.11 | Morbid obesity | endocrine/metabolic | 0.083 | 1.087 | 0.020 | 3.00E-05 | 467399 | 8411 |
